# Brain Ageing in Social Anxiety Disorder: An ENIGMA-Anxiety Mega-Analysis Across 26 International Cohorts

**DOI:** 10.64898/2026.07.02.26357108

**Authors:** Kimberly V. Blake, Jonathan C. Ipser, Alyssa R. Amod, Tobias Kaufmann, Yair Bar-Haim, Jochen Bauer, Ali Bayram, Katja Beesdo-Baum, Laura Blanco-Hinojo, Tiana Borgers, Robin Bülow, Marta Cano, Narcís Cardoner, Christopher R. K. Ching, Soo-Hee Choi, Udo Dannlowski, Christopher G. Davey, Alexander G. G. Doruyter, Kira Flinkenflügel, Gregory A. Fonzo, Tomas Furmark, Dominik Grotegerd, Hans J. Grabe, Tim Hahn, Ben J. Harrison, Alexandre Heeren, Kevin Hilbert, Yoshiyuki Hirano, Joy Hirsch, David Hofmann, Yuko Isobe, Neda Jahanshad, Hamidreza Jamalabadi, Alec J. Jamieson, Andreas Jansen, Jaehyun Edmund Kim, Tilo Kircher, Hitomi Kitagawa, Anna Luisa Klahn, Saskia B. J. Koch, Axel Krug, Harald Kugel, Dasom Lee, Elisabeth J. Leehr, Christine Lochner, Ulrike Lueken, Amirhossein Manzouri, Kristoffer N. T. Månsson, Koji Matsumoto, Susanne Meinert, Alicia Menze, Markus Muehlhan, Benson Mwangi, Igor Nenadić, Ziphozihle Ntwatwa, Hyuntaek Oh, Spiro P. Pantazatos, Martin P. Paulus, Jutta Peterburs, Jesus Pujol, Karin Roelofs, Ramiro Salas, Franklin R. Schneier, Elisabeth Schrammen, Eiji Shimizu, Lisa Sindermann, Theresa M. Slump, Jair C. Soares, Benjamin Straube, Thomas Straube, Murray B. Stein, Ardesheer Talati, Florian Thomas-Odenthal, Sophia I. Thomopoulos, Raşit Tükel, Anna Tyborowska, Marie-José Van Tol, Dick J. Veltman, Roman A. Vogler, Inge Volman, Henry Völzke, P. Michiel Westenberg, Katharina Wittfeld, Mon-Ju Wu, Noga Yair, Tokiko Yoshida, Chen Zhang, Xi Zhu, Giovana B. Zunta-Soares, Peter Zwanzger, Daniel S. Pine, Moji Aghajani, Paul M. Thompson, Nic J. A. Van der Wee, Dan J. Stein, Janna Marie Bas-Hoogendam, Nynke A. Groenewold

## Abstract

Social anxiety disorder (SAD) is among the most prevalent anxiety disorders, and it has been associated with signs of advanced biological ageing. Despite this, brain age research on anxiety disorders remains limited. This mega-analysis investigated brain ageing in adults with SAD within the ENIGMA-Anxiety Working Group. Structural MRI scans from 576 participants with SAD and 1 355 non-affected healthy controls (HCs) across 26 international samples were included. Brain age was estimated from 77 cortical and subcortical regions using a publicly available ENIGMA brain age model. The brain-predicted age difference (brain-PAD) was calculated as the difference between brain age and chronological age. Group and subgroup differences (comorbidity, medication) were assessed using linear mixed-effect models. In the full sample, there was no group difference in brain-PAD (**β**diagnosis (SE)=0.70 (0.37) years, *p*=0.061). In a subgroup of participants with SAD with comorbid anxiety disorders (n=184 SAD, n=1 355 HCs), a brain-PAD of +2.39 (0.93) years (Cohen’s *d*=0.23, *p*FDR=0.003) was observed. This brain-PAD became smaller after exclusion of participants with comorbid agoraphobia and specific phobia, suggesting that these disorders may partly drive the advanced brain-PAD. In conclusion, this ENIGMA-Anxiety mega-analysis did not find evidence of advanced brain ageing in the full sample of adult participants with SAD relative to HCs. However, a sub-analysis suggested that SAD with co-occurring phobic disorders, or the phobic disorders themselves, are associated with neurostructural patterns typical of older brains. Future research could utilise transdiagnostic samples with information on age of onset and disorder duration to further clarify this relation.

## Introduction

Social anxiety disorder (SAD) is defined by a marked fear of one or more social situations and the fear of negative evaluation from others (1). It is one of the most common anxiety disorders, with lifetime prevalence ranging from 0.2%-12.1% across countries (2; 3). Cross-national data demonstrate that SAD is an international public health concern due to its high prevalence and debilitating nature (3–4). Further, SAD seldom exists in isolation, frequently presenting with comorbid psychopathology, including additional anxiety and mood disorders (2; 5). As a result, anxiety disorders have been associated with an early mortality or increased risk of disability in older age (6). Indeed, individuals with anxiety disorders may have a greater risk of developing age-related medical conditions, such as cardiovascular conditions (7). Emerging evidence furthermore suggests that structural brain abnormalities in SAD, such as less grey matter (GM), and white matter (WM) abnormalities, may be associated with advanced biological ageing (8; 9). Therefore, investigating brain ageing in participants with SAD may provide critical insight into the neurobiological mechanisms linking anxiety and brain ageing.

In previous work, SAD has been associated with alterations in brain morphology (10–14). The largest multisite, cross-national mega-analysis to date, performed by the ENIGMA-Anxiety (Enhancing NeuroImaging Genetics through Meta-Analysis) Working Group (15) on participants with SAD (n=1 115) and non-affected healthy controls (HCs; n=2 775; 11), demonstrated smaller bilateral putamen volume in participants with SAD. There was also a significant interaction between SAD and age in the left putamen, with adult, but not adolescent, participants with SAD showing smaller bilateral putamen volumes and larger bilateral pallidum volumes compared to HCs (11).

A promising method for investigating advanced biological ageing in SAD involves the application of machine learning (ML) models of brain age. Brain age ML models are typically trained on large datasets, and then applied to the neuroimaging data of an individual, such as structural or functional magnetic resonance imaging (MRI), to make age predictions (16–19). The deviation between estimated brain age and chronological age is the brain-predicted age difference (brain-PAD). A positive brain-PAD indicates that an individual’s brain appears “older” than would be predicted based on their chronological age (19). The brain age method can be used to develop a reference model derived from healthy individuals, and subsequently to identify structural brain ageing patterns that may be associated with disease (20–21).

The brain-PAD has become an increasingly popular tool for investigating risk for atypical brain ageing in psychiatric disorders: this includes psychotic, mood, and anxiety disorders, which have shown higher brain-PAD in comparison to HCs (22-23; 19; 24-27). Moreover, higher brain-PAD seems to be associated with more severe symptomology in mood and psychotic disorders (24; 28-30). Of note, lower brain-PAD has been reported in psychotropic medication-treated compared with untreated participants with major depressive disorder (MDD) and bipolar disorder, which may reflect neuroprotective effects of these agents (24; 31).

Only three studies have investigated brain ageing in adults with anxiety disorders. The first examined participants with anxiety disorders (18-57 years; generalised anxiety disorder, panic disorder, or SAD; n=67) compared to HCs (n=65) and found that when including antidepressant use as a covariate, anxiety patients had a brain-PAD of +2.91 years compared to HCs (24). The second study, conducted within the ENIGMA-Anxiety Working Group, included a large sample of participants with specific phobia (SPH; n=600) compared to HCs (n=1 134), aged 22-75 years, from 17 sites (32) and reported no group difference in brain-PAD between participants with SPH and HCs. However, a robust diagnosis-by-age interaction was identified across analyses, suggesting subtle, advanced brain ageing in younger formally diagnosed participants with SPH. The third study, conducted within the ENIGMA-Anxiety Working Group, investigated brain ageing in both children and adults with generalised anxiety disorder (GAD), finding no group difference between GAD and HCs. However, the study found a positive relationship between symptom severity and brain-PAD (33). As the study by Han et al. (24) was a transdiagnostic study on a single-site clinical sample, and the studies by Blake et al., (32) and Richier et al. (33) investigated brain-PAD in adults diagnosed with SPH, and GAD, respectively, it is apparent that there is a gap in the literature regarding brain ageing in a large sample of adults with SAD.

The first and primary aim of the present study was to compare brain-PAD in adults with SAD versus HCs, in a large, cross-national cohort. We hypothesised that adults with SAD would have a higher brain-PAD than the HC group. Furthermore, secondary analyses investigated differences in brain-PAD in subgroups of SAD compared to HCs, as defined by comorbidity and medication status. The final aim of the study was to test the hypothesise that SAD symptom severity would be positively associated with brain-PAD.

## Methods

### Study Sample

This multi-site, cross-national ENIGMA-Anxiety mega-analysis included an initial sample of adult participants diagnosed with SAD (n=632) and HCs (n=1 667) from 26 study sites, aged 22-68 years. The study sites, from 10 countries, collected demographic and clinical information for each participant as summarised in Table 1a, b. As reported in Groenewold et al. (11), participants with SAD were excluded if they had a lifetime diagnosis of bipolar disorder, schizophrenia spectrum disorder, or autism spectrum disorder. Participants with SAD with other comorbidities, including other anxiety disorders, MDD, and substance use disorder, were retained, however. HCs were excluded if they had a current or lifetime major psychiatric diagnosis or psychotropic medication use at the time of the scan.

**Table 1a.**
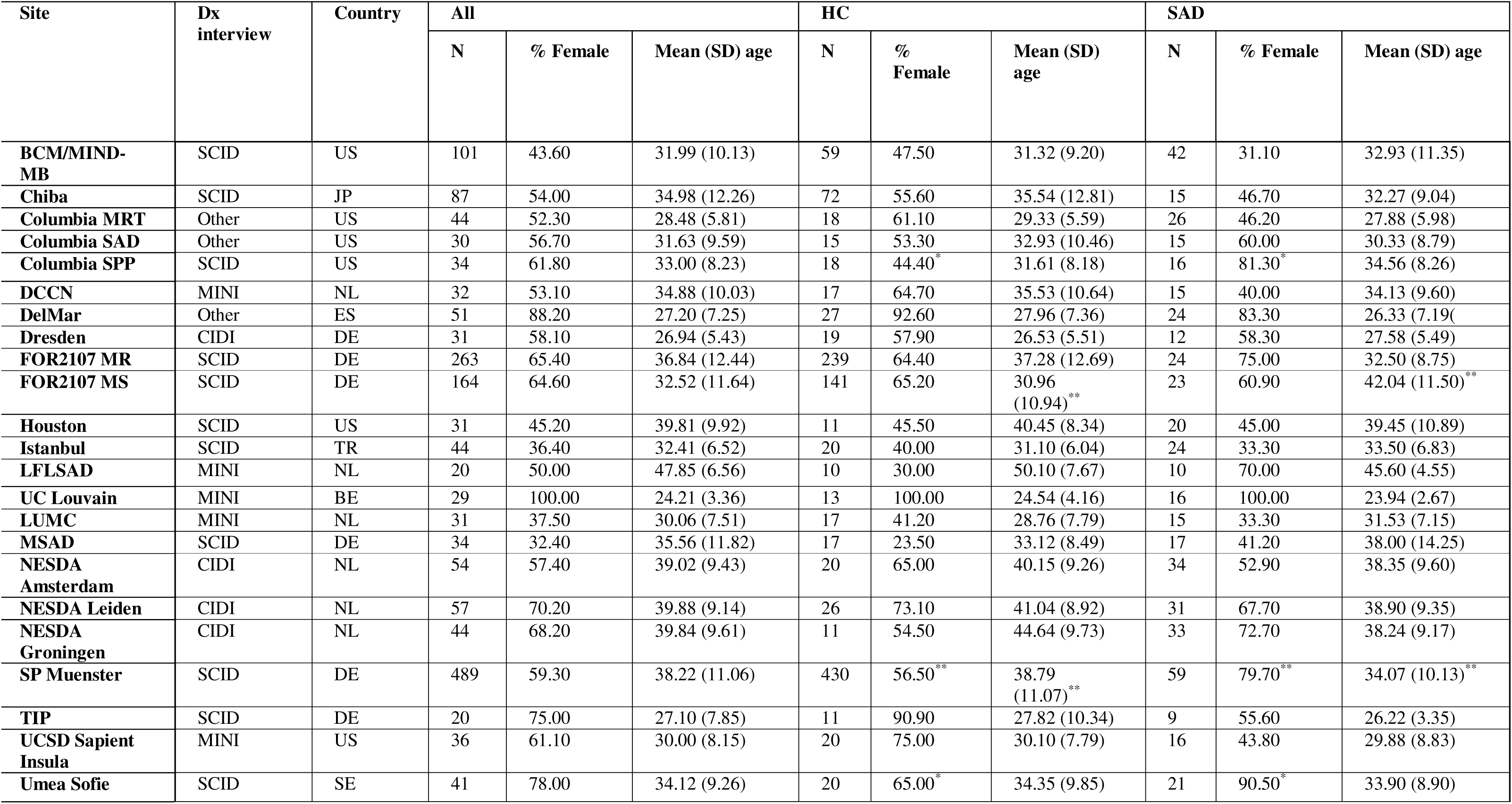
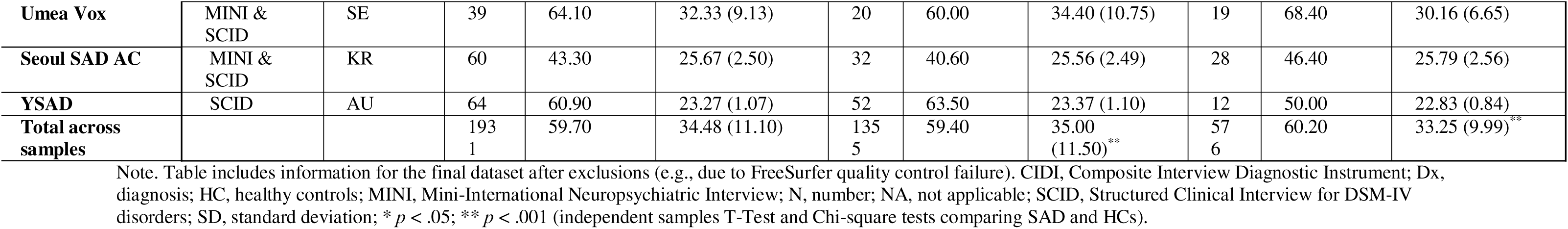
Sociodemographic information for each study site included in the main analyses.

**Table 1b.**
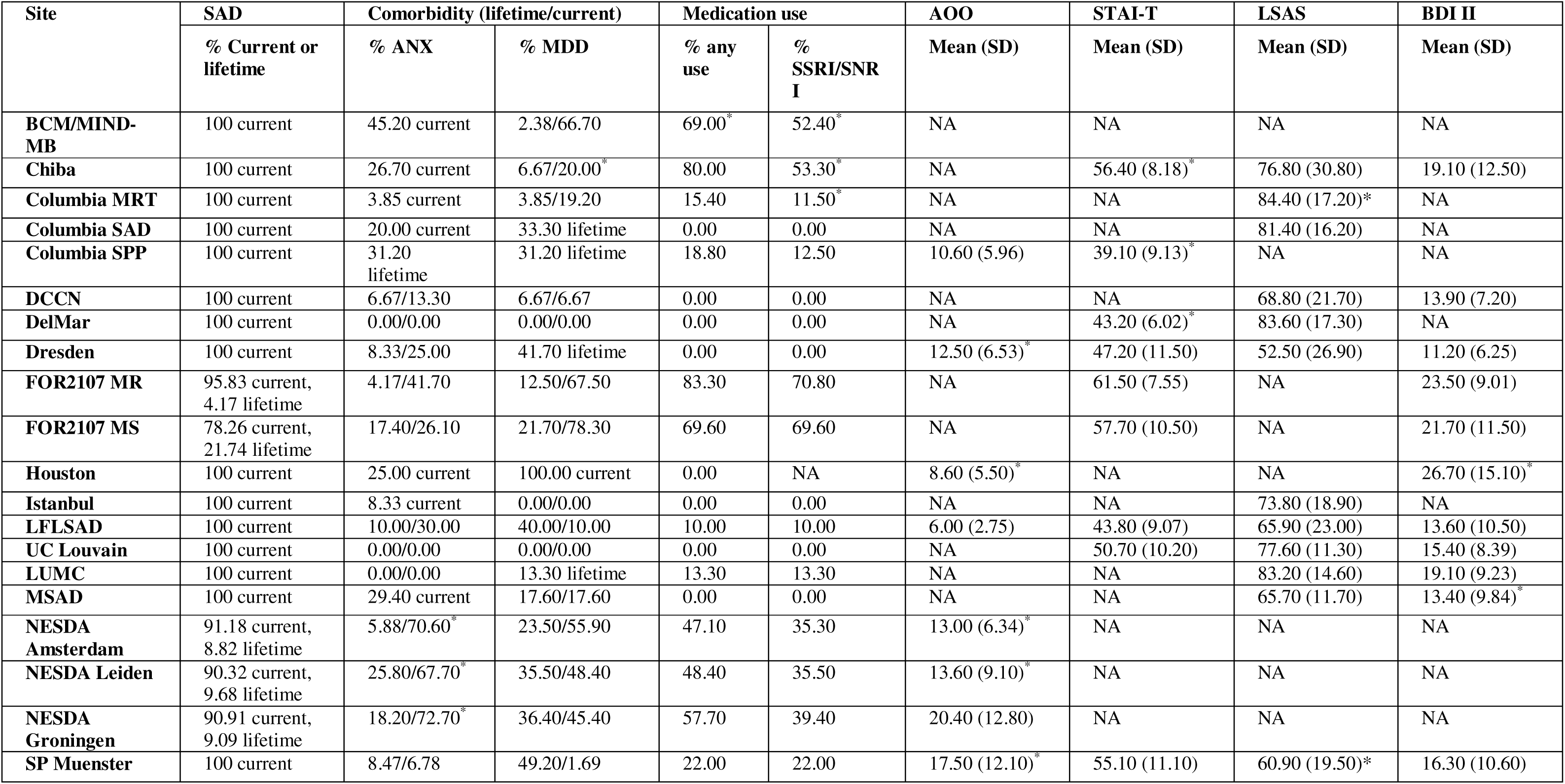
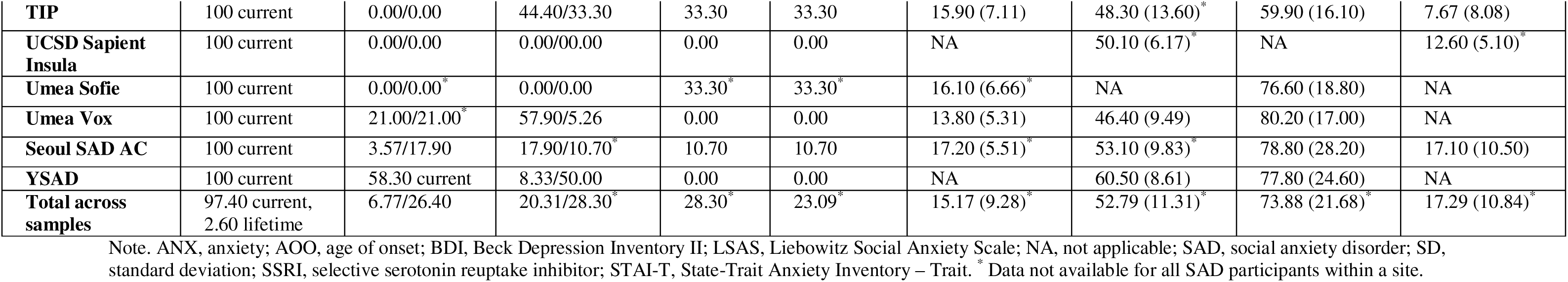
SAD clinical information for each study site included in the main analyses.

The Human Research Ethics Committee (HREC) at the University of Cape Town granted approval for this study (HREC reference: 675/2021). Each study site obtained informed participant consent and approval from ethical and institutional review boards prior to data collection, and consent for data sharing within the ENIGMA-Anxiety Working Group.

### Image Acquisition and Processing

At the original study sites, investigators processed 3D volumetric structural T1-weighted brain MRI scans using FreeSurfer (version 5.1-7.4.1, with 5.3 being the most commonly used version; 34) and generated FreeSurfer output summary statistics and histograms for visual inspection. FreeSurfer output then underwent established ENIGMA quality control (QC) protocols (https://github.com/ENIGMA-git/ENIGMA-FreeSurfer-protocol; 35-36) by the investigators at the original sites and investigators of the current project. FreeSurfer regions were excluded due to segmentation failure, poor quality segmentations due to processing errors or severe global misclassifications, over/underestimations and misclassifications of brain regions, or motion artifacts. Participants with > 10% missing FreeSurfer data were removed from analyses (SAD n=56, HC n=118). In addition, HCs who were part of a previous ENIGMA-MDD brain age model training were excluded (19; n=194), resulting in a final sample of 576 participants with SAD (lifetime n=15, current n=561) and 1 355 HC (supplementary Figure S1: age density plot).

### Brain Age Model

A ridge regression model developed by the ENIGMA-MDD Working Group (19) was used to predict brain age on the merged dataset of the current project. This brain age model was developed by creating normative models of the association between 77 structural brain features and chronological age in the training control group using the Python-based *sklearn* package version 0.23.1 (37). The model estimated brain age by combining FreeSurfer measures (34) from the right and left hemispheres and calculating the mean volumes of lateral ventricles and subcortical volumes, and cortical thickness and surface area based on the Desikan/Killiany atlas (38). The ENIGMA-MDD model was trained separately on control females (n=14 182) and males (n=12 353), age range: 18-75 years (19). Model validation in a test set of HCs had a mean absolute error (MAE) of 6.50 (SD=4.91) years in male HCs and 6.84 (SD=5.32) years in female HCs. The Pearson correlation between brain age and chronological age was r=0.85 for the male model, and 0.83 in the female models (both *p*<.001). The ENIGMA-MDD brain age model was selected because of its comparable sample characteristics and alignment in data processing pipelines.

### Statistical Analyses

The present study followed a mega-analytic framework as evidence suggests its superiority compared to meta-analysis regarding data preservation, statistical power and flexibility in covariate adjustment (39–41). Brain age estimates were obtained on each site’s FreeSurfer data on a local PC with Python (version 3.6.2), following the methods described in Han et al. (19). The brain-PAD was calculated by subtracting each participant’s chronological age from their brain age.

The fit of the ENIGMA-MDD model with the current study data was assessed by calculating the MAE and Pearson correlation coefficients and explained variance (R2) between brain age and chronological age in the whole sample, and separately for diagnostic groups (SAD; HC) and by sex. Model fit statistics were inspected for each research site. The dataset was checked for normality for brain-PAD, MAE, chronological age and cortical thickness (supplementary Figure S2a-S5b). Outliers were identified based as >1.5 x IQR beyond the quartiles, and extreme outliers as >3 x IQR beyond the quartiles.

Using the *nlme* package (version 3.1-162) in RStudio, a linear mixed-effects model (LME) was used for the first primary analysis to test for a potential group difference in brain-PAD. Brain-PAD was included as the outcome variable (unstandardised **β**-values, measured in years) and diagnosis (SAD; HC) as the predictor variable. To mitigate potential age bias, mean-centred chronological age and mean-centred chronological age squared were included as covariates, in addition to sex. Scan site was included in the models as a random intercept. Diagnosis-by-sex, diagnosis-by-chronological-age (linear) and diagnosis-by-chronological-age-squared (quadratic) interaction terms were included in the model, one at a time, to evaluate if the relationship between the brain-PAD, chronological age and sex differed by group. Interaction variables were removed from the model if they were not significant, to retain best model fit. Bayesian information criterion (BIC) and Akaike information criterion (AIC) determined which model had the best fit (40).

Mean chronological age of the SAD and HC groups differed significantly for two scan sites (FOR2107MS SAD = 30.96 years, HC = 42.04 years; SP Muenster SAD = 38.79 years, HC = 34.07 years; Table 1a). Using the *MatchIt* R package (version 4.5.0), we conducted propensity score matching to match groups (SAD/HC) within these sites on age. Nearest neighbour matching was selected as this approach selects the closest eligible control participant for each case (42). We matched two HCs to one SAD participant on age to optimize sample size, and the distance measure used was the difference between the propensity scores of each case and control unit. The primary LME was rerun in this propensity score matched sample.

A sensitivity analysis was conducted in participants with current SAD only compared to HCs. Comorbidity subgroup analyses were conducted, repeating the model in participants with SAD, with and without comorbid MDD, and in participants with SAD, with and without anxiety disorder comorbidities (GAD, SPH, panic disorder (PD), agoraphobia (AG), separation anxiety, and unspecified anxiety disorder). In addition, medication subgroup analyses were conducted comparing the brain-PAD between HCs and unmedicated SAD, and between HCs and medicated SAD. False discovery rate (FDR) correction was applied to these six subgroup analyses, with a significance threshold of *p*<0.05.

Finally, several post-hoc sensitivity analyses were conducted. First, the primary model was rerun after excluding data from three NESDA sites (SAD n=105, HC n=1,298), as these sites appeared to have lower cortical thickness values than other sites. Next, the primary model was rerun in the SAD group with anxiety disorder comorbidities, compared to HCs, this time removing each anxiety disorder comorbidity subgroup (GAD, PD, AG and SPH) from the entire sample, one at a time.

To address the last aim of the study, analyses were run to determine whether an association between brain ageing and SAD symptom severity was present. For these analyses, data from the Liebowitz Social Anxiety Scale (LSAS; 43), measuring symptom severity in relation to specific social situations, and the trait version of the State Trait Anxiety Inventory (STAI-T; 44), measuring trait anxiety, were selected. A mixed-effects, random intercept regression model was conducted in participants with SAD only (no HCs), with brain-PAD as the outcome variable, LSAS score as the predictor variable and mean-centred chronological age, mean-centred chronological age squared, and sex as covariates. The analysis was repeated with STAI-trait score as the predictor variable. All analyses were conducted in R (version 4.2.1).

## Results

### Model Fit

The brain age model fit with this multi-site ENIGMA-Anxiety SAD dataset was comparable to that seen in prior studies (45; 19; supplementary Table S1). Model fit statistics in the present study were as follows: MAE full sample=6.90 (SD=5.32) years, SAD=7.53 (5.81), HC=6.62 (5.07). When weighted by sample age range (MAE divided by sample age range), the weighted MAE was as follows: full sample=0.15; SAD=0.16; HC=0.14. Finally, Pearson correlations between brain age and chronological age were as follows: full sample=0.69, SAD=0.65, HC=0.72 (supplementary Table S2 and S3: model fit per study site).

### First Aim: Brain-PAD in SAD Versus HCs

Participants with SAD (n=576) had a mean chronological age of 33.25 (SD: ± 9.99), brain age of 36.64 (SD: ± 11.05) years, and mean brain-PAD of 3.39 (SD: ± 8.89) years. HCs (n=1355) had a mean chronological age of 35.00 (SD: ± 11.50), brain age of 35.77 (SD: ± 10.44) years and mean brain-PAD of 0.76 (SD: ± 8.31) years. After adjusting for scanner site, mean-centred chronological age, mean-centred chronological age squared and sex, brain-PAD did not differ significantly between participants with SAD and HCs. There was, however, a trend towards a significant difference (*p*=0.061, Table 2; Cohen’s *d* (95% confidence intervals) = 0.08 (-0.01-0.17), (Figure 1a; supplementary Figure S6: scatterplots per site; supplementary Figure S7: brain-PAD residuals). A significant main effect of mean-centred chronological age and mean-centred chronological age squared was present (Table 2). Data from one participant with SAD and one HC were identified as outliers due to extreme positive and negative brain-PADs in SPSS (version 30, supplementary Table S4, Figure S8). As the outliers did not significantly impact model output, they were retained (supplementary Table S5 for model output).

**Figure 1.**
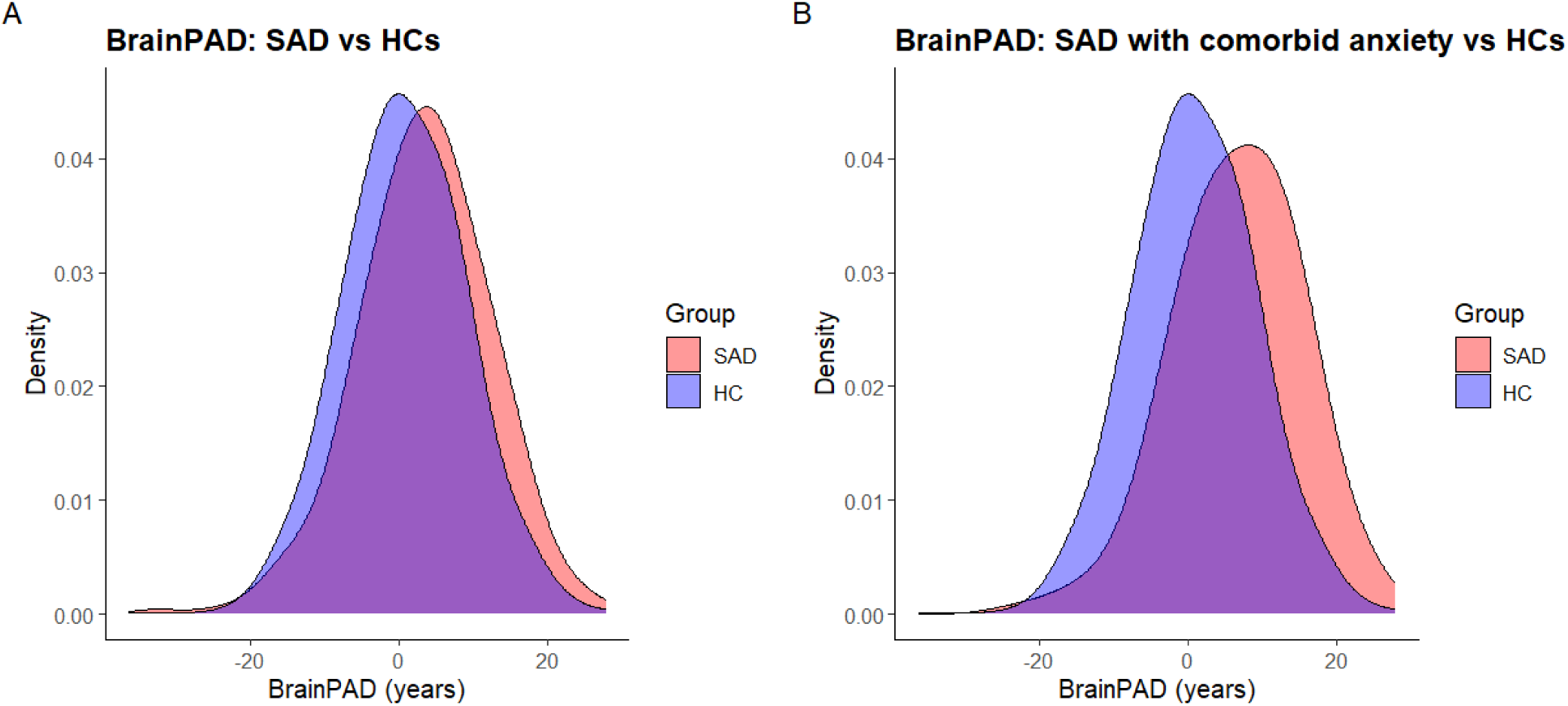
a. Density plot for brain-PAD in SAD and HCs in the full sample. Figure 1b. Density plot for subgroup of SAD with anxiety disorder comorbidities and HCs. Note. HC, healthy controls; SAD, social anxiety disorder.

**Table 2.** Between-group differences (SAD vs HC) in brain-PAD.

| | $\beta$ (SE) | t-value | p |
| --- | --- | --- | --- |
| **Intercept (site)** | 2.24 (0.96) | 2.33 | **0.020** |
| **Diagnosis** | 0.70 (0.37) | 1.88 | 0.061 |
| **Sex** | -0.18 (0.32) | -0.56 | 0.577 |
| **AgeC** | -0.32 (0.02) | -16.29 | **<0.001** |
| **AgeC<sup>2</sup>** | -4.34 x 10<sup>-3</sup> (1.47 x 10<sup>-3</sup>) | -2.96 | **0.003** |

Note. AgeC, mean-centered chronological age; AgeC<sup>2</sup>, mean-centered chronological age squared; HC, healthy controls; SAD, social anxiety disorder; SE, standard error. $\beta$ is measured in years.

Interaction terms did not contribute significantly to the model (**β**diagnosis-by-chronological-age=-0.04 (0.03), t-value=-1.08, *p*=0.282; **β**diagnosis-by-chronological-age-squared=-0.01 (0.00), t-value=-1.85, *p*=0.064; **β**diagnosis-by-sex=-0.58 (0.69), t-value=-0.83, *p*=0.404). In certain model combinations where diagnosis-by-chronological-age-squared was included, the group difference in brain-PAD reached significance. However, as the interaction terms did not significantly contribute to the model, they were removed and were treated as supplemental (supplementary Table S6: models including interactions). The model without interaction terms and with the best fit was retained (Table 2), as determined by lower AIC (=1 292.27) and BIC (=1 2981.21).

### Planned Sensitivity Analyses: SAD Versus HCs

The primary LME was rerun in participants with current SAD (removing those with lifetime, but not current, diagnosis; remaining n=561), compared to HCs. The group difference in brain-PAD was similar, as expected, approaching significance (**β**=0.72 (0.38), *p*=0.056, supplementary Table S7 for full model results).

Finally, the model was rerun in the dataset where data from two sites were propensity score matched on chronological age (supplementary Table S8, Figure S9). In this dataset, the brain-PAD was not significantly different between participants with SAD and HCs, and the **β**-value was slightly smaller in size (**β**=0.69(0.39), *p*=0.07; supplementary Table S9).

### Secondary Clinical Subgroup Analyses

Next, we ran the LME in participants with SAD with either current or lifetime comorbid anxiety (PD, GAD, SPH, AG and mixed anxiety-depressive disorder), compared to HCs. A significant brain-PAD of +2.39 (0.93), Cohen’s *d*=0.23, *p*FDR<0.003 was present between the subset of participants with SAD with comorbid anxiety (n=184) compared to HCs (n=1 355, Table 3, Figure 1b; supplementary Table S10, S11 and S12 for subgroup clinical characteristics). However, when we compared participants with SAD with MDD comorbidity to HCs, no group difference in brain-PAD was present (**β**=0.75 (0.49), *p*FDR=0.208, Table 3).

**Table 3.** Between group differences in brain-PAD, SAD clinical subgroups compared to HCs.

|  | SAD | HC | Dx |  |  |  |  |
| --- | --- | --- | --- | --- | --- | --- | --- |
| | n | n | $\beta$ (SE) | t-value | p | pFDR | Cohen's d (95% CI) |
| SAD (medication use) vs HCs | 163 | 1 355 | 1.18(0.60) | 1.96 | 0.051 | 0.152 | 0.14(0.04-0.24) |
| SAD (ANX comorbidity) vs HCs | 184 | 1 355 | 2.39(0.93) | 2.58 | **<0.001** | **0.003** | 0.23(0.13-0.33) |
| SAD (MDD comorbidity) vs HCs | 280 | 1 355 | 0.75(0.49) | 1.55 | 0.121 | 0.208 | 0.08(-0.01-0.18) |
| SAD (no medication use) vs HCs | 399 | 1 355 | 0.49(0.44) | 1.10 | 0.271 | 0.326 | 0.05(-0.04-0.15) |
| SAD (no ANX comorbidity) vs HCs | 392 | 1 355 | 0.10(0.42) | 0.22 | 0.823 | 0.823 | 0.01(-0.08-0.10) |
| SAD (no MDD comorbidity) vs HCs | 296 | 1 355 | 0.72(0.49) | 1.48 | 0.139 | 0.208 | 0.08(-0.02-0.18) |

Note. ANX, anxiety; CI, confidence intervals; Dx, diagnosis; FDR, false discovery rate; HC, healthy controls; n, number; SAD, social anxiety disorder; SE, standard error. $\beta$ is measured in years.

Finally, the LME was run in participants with SAD taking medication at the time of the scan compared to HCs. Here, the diagnosis **β**-value was larger than in the full dataset (**β**=1.18 (0.60)), and the group difference in brain-PAD was borderline significant (*p*=0.051, Table 3), however, this did not survive correction (*p*FDR=0.152). The LMEs run in participants with SAD without medication use, in participants with SAD without anxiety disorder comorbidities, and in participants with SAD without MDD comorbidity (all models: compared to HC) also did not reveal group differences in observed brain-PAD (Table 3).

### Post-Hoc Sensitivity Analyses: Clinical Subgroups

To follow up on the significant group difference in the subset of 184 participants with SAD with comorbid anxiety, we ran multiple sensitivity analyses. A previous ENIGMA study demonstrated that NESDA participants with MDD and HCs have lower average cortical thickness, and therefore overestimated brain age (24). The NESDA study also has high levels of anxiety comorbidity (comorbid MDD and anxiety = 41.4%). Therefore, the LME was repeated after removal of the three NESDA sites. The group difference held (**β**=1.92 (0.70), *p*=0.006, supplementary Table S13 for full model results).

To determine whether any one anxiety disorder comorbidity (GAD, PD, AG, or SPH) was driving the effect seen, a post-hoc sensitivity analysis was run in the SAD group with anxiety comorbidities compared to HCs, removing participants with each comorbidity (GAD, PD, AG and SPH) one diagnosis at a time. All group differences in brain-PAD remained significant, but effect sizes were lower in LMEs after removal of comorbid AG and SPH (supplementary Table S14).

### Second Aim: Symptom Severity

LSAS and STAI-T scores were not significantly associated with brain-PAD in participants with SAD (LSAS score: n=318, **β**LSAS=0.02 (0.02), t-value=0.78, *p*=0.434; STAI-T score: n=248, **β**STAI-T=0.04 (0.04), t-value=0.88, *p*=0.378).

## Discussion

This ENIGMA-Anxiety mega-analysis investigated whether adult participants with SAD showed a larger brain-PAD than HCs. No group difference in brain-PAD was present in the full sample (*p*=0.061) and the effect size was relatively small (Cohen’s *d*=0.08). Secondly, the brain-PAD did not relate to SAD symptom severity, as measured with the LSAS, or trait anxiety, as measured with the STAI-T. Secondary analyses in clinical subgroups did reveal an effect, however, with a positive brain-PAD of +2.39 (0.93) years (Cohen’s *d*=0.23) present in a clinical subgroup of participants with SAD with comorbid anxiety disorders, compared to HCs, after adjusting for chronological age, sex, and study site (*p*FDR=0.003). Post-hoc sensitivity analyses revealed that this group difference was not dependent on a specific comorbid anxiety disorder, as the effect remained significant when data from participants with SAD with specific comorbid anxiety disorders were removed one at a time. In addition, the brain-PAD became smaller after exclusion of participants with comorbid AG and SPH. These findings indicate that SAD with comorbid anxiety disorders may be related to advanced brain ageing, or alternatively, that the comorbid conditions are associated with advanced brain ageing, rather than the SAD itself.

### Brain Ageing in Social Anxiety Disorder

While several studies have reported advanced brain ageing in mood and psychotic disorders (22; 19; 24-25), the present mega-analysis did not identify a group difference in brain-PAD in the full sample of participants with SAD and HCs. Notably, the group difference was close to significance, and when certain interaction variables were included in the model (e.g. diagnosis-by-chronological-age-squared and diagnosis-by-sex; supplementary Table S7), the difference reached significance. However, as these interactions did not significantly contribute to the regression model, they were removed to retain optimal model fit. In a sensitivity analysis including only SAD with current diagnosis (not lifetime) compared to HCs (n=561), the group difference was again just short of significance, with a slightly larger **β**-value than in the full sample (**β**=+0.72 years (0.38), Cohen’s *d*=0.08, *p*=0.056; supplementary Table S13).

An important consideration in brain age research is potential bias related to chronological age. Age is often underestimated in older participants and overestimated in younger participants by brain age models (46). We attempted to manage this bias by rerunning the primary LME in a dataset that was propensity score matched for age between cases and HCs, with the group difference in brain-PAD still not achieving statistical significance. Further, the present study controlled for age by including this variable as a linear and quadratic term in the LMEs (47–48), therefore mitigating age bias.

### Clinical Subgroup Findings

Notably, a positive brain-PAD of +2.39 years was identified in participants with SAD with a comorbid anxiety disorder (GAD, PD, AG, SPH, other), compared to HCs. In contrast, there was no group difference in brain-PAD in participants with SAD without anxiety comorbidities, compared to HCs. The brain-PAD in participants with SAD with comorbid anxiety remained significant in several sensitivity analyses, but when removing participants with phobic disorders (AG and SPH), we observed a decrease in brain-PAD **β**-value by approximately 1 year, indicating that AG and SPH may partially drive the group difference (SAD no AG: **β**=1.72 years (0.72), Cohen’s *d*=0.20, *p*=0.017; SAD no SPH: **β**=1.72 years (0.66), Cohen’s *d*=0.18, *p*=0.009). Sample sizes after removal of comorbid AG and SPH were slightly higher than after removal of comorbid GAD and PD, potentially impacting statistical power (SAD no GAD: **β**=2.88 years (0.76), Cohen’s *d*=0.33, *p*<0.001; SAD no PD: **β**=2.65 years (0.73), Cohen’s *d*=0.32, *p*<0.001; supplementary Table S14). Interestingly, SPH may be considered a marker for subsequent internalising disorders, in part due to its particularly early age of onset during childhood (49–50). These findings underscore the importance of assessing comorbid anxiety presentations in clinical settings, as they may reflect more advanced neurostructural brain ageing.

The size of the present brain-PAD estimate in participants with SAD with anxiety disorder comorbidities (+2.39 years) is smaller than that observed in psychotic disorders (+3-4 years), similar to bipolar disorder (+2 years; 51; 22; 25), and larger than MDD (+1 year; 19). The brain-PAD identified in the present study is comparable in **β**-value and effect size (Cohen’s *d*=0.23) to that found in a cross-disorder sample of participants with anxiety disorders (SAD, GAD, PD) compared to HCs, after correction for antidepressant use (+2.91 years Cohen’s *d*=0.27; 24). The similarities in effect size across studies support the utility of brain-PAD as an index of neurostructural deviations in anxiety disorders.

Multiple biological variables could contribute to advanced brain ageing, such as that observed in the clinical subgroup of participants with SAD with anxiety comorbidities. Oxidative stress may be a major component of anxiety pathology (52), and has been implicated in premature cell death and biological ageing (53–54). In addition, human and animal studies suggest that stress-induced inflammation may harm brain function, in turn, negatively impacting mental health and potentially contributing to the aetiology of anxiety disorders (55).

The positive brain-PAD observed in participants with SAD and anxiety comorbidities may suggest that individuals with multiple anxiety disorders, as opposed to one, exhibit more pervasive impairment (56), although functional impairment was not measured in the present study, and participants with different combinations of comorbidities were not directly compared due to small sample sizes. While we did not see higher levels of symptom severity (LSAS, STAI-T) in the SAD group with anxiety comorbidities (supplementary Table S12) than in other clinical subgroups, these measures may not adequately capture longer term clinical course. Research has shown that patients with multiple anxiety disorders report higher symptom severity than those with one anxiety disorder (57–58), and that long term disability was highest in participants with SAD and multiple anxiety comorbidities, as opposed to GAD only, PD with AG, and PD only (59). Moreover, a study that compared the effect of anxiety disorder combinations in 1 004 outpatients found that level of functioning declined as number of comorbidities increased, and the combination of SAD with PD was the most debilitating (56).

Due to potential subtle neuroprotective effects of psychotropic medication use on brain ageing (51), and because anxiety disorders frequently present with comorbidities (5), participants were further split into clinical subgroups based on medication use and comorbidity with MDD. Contrary to previous brain age literature (24; 31), we found a positive brain-PAD of approximately 1 year in participants with SAD taking psychotropic medication, compared to HCs. However, this difference was not significant after correction for multiple comparisons (*p*FDR = 0.152). Approximately 44% of the clinical subgroup of participants with SAD with anxiety comorbidities took medication at the time of the scan, and close to 77% had comorbid MDD (Supplementary Table S12). However, no group difference in brain-PAD was present in the subgroup of participants with SAD with MDD (50.36% of whom also had comorbid anxiety disorders), compared to HCs, indicating that MDD did not explain the group difference. This could potentially be due to anxiety disorders being the primary diagnoses in our sample, however, this information was not available and therefore could not be formally assessed.

### Social Anxiety Symptom Severity and Trait Anxiety

Our second research aim was to investigate whether there was an association between brain-PAD and symptom severity in participants with SAD. Contrary to previous studies that identified a positive association between the brain-PAD and symptom severity in mood, anxiety and psychotic disorders (24; 28-30), neither metric was associated with brain-PAD in the present study. Another ENIGMA-Anxiety mega-analysis, from which the present sample was derived, also found no associations between subcortical volumes and LSAS and STAI-T score (11). While the LSAS is a well-validated and reliable measure of social anxiety symptoms, its reliance on self-report may lead to under- or overreporting of symptoms (60). In addition, the LSAS is designed to capture current or recent social anxiety severity, as opposed to longer-term severity (43). These measures therefore may not capture additional clinically relevant variables, such as disorder chronicity. Despite our null findings, our results in subgroups of participants with SAD with comorbid anxiety suggest that exploring such clinical characteristics may provide further insight into brain age in anxiety disorders.

### Study Strengths, Limitations and Future Recommendations

This study is the first mega-analysis to investigate brain ageing in a large, cross-national sample of adult participants with SAD. The brain age model showed a moderate fit with the present sample. Further, the mega-analytic design of the present ENIGMA-Anxiety study enabled the use of pre-established preprocessing pipelines and uniform handling of covariates, thereby reducing heterogeneity (61; 41).

Although mega-analyses allow for larger sample sizes, limited power may still be present in this study, particularly for differences of less than one year. Multiple group differences in brain-PAD were close to the significance threshold of *p*=0.05, and a subtle, positive **β**-value of +0.70 years was present in the full sample of participants with SAD. This suggests that an increase in sample size, using a brain age model with lower MAE, or closer harmonisation of neuroimaging data across sites, may have resulted in these group differences reaching significance. An additional limitation of this study is its cross-sectional design, which restricts the ability to infer causal relationships (62; 63). Further, not all sites assessed phobic disorders, therefore it is possible these disorders remained present in the HC group. Finally, the present study was unable to control for certain potentially confounding factors that may impact brain age, such as heavy alcohol consumption and higher blood pressure (64), as data on these lifestyle variables were not consistently available.

Future studies should use more sophisticated brain age models, such as deep learning-based brain age models (65). Further, large-scale transdiagnostic studies with larger clinical subgroup samples may help elucidate how comorbid anxiety disorders, or specific combinations of anxiety disorders, relate to brain ageing.

## Conclusions

The present ENIGMA-Anxiety mega-analysis did not find evidence of advanced brain ageing in the full sample of participants with SAD and HCs. Additionally, no associations between brain-PAD and SAD symptom severity or trait anxiety were present. However, an advanced brain-PAD of +2.39 years was identified in participants with SAD with comorbid anxiety disorders. This effect was smaller following the exclusion of participants with comorbid AG and SPH, indicating that co-occurring phobic disorders may partly drive the advanced brain-PAD observed in this subgroup. Taken together, these findings suggest that SAD comorbid with anxiety disorders, particularly AG and SPH, may reflect greater neurostructural alterations, represented by advanced brain ageing. This implies that these individuals exhibit neurostructural alterations typically observed in older brains. Future work utilising transdiagnostic approaches may benefit from integrating clinical information, such as age of onset and disorder duration, with biological and behavioural data, to further uncover the nuances of this relationship.

## Data Availability

The ENIGMA-Anxiety Working Group is open to sharing the data and code from this
investigation to researchers for secondary data analysis. To request access to brain morphometric, clinical, and demographic data, an analysis plan can be submitted to the ENIGMA-Anxiety Working Group (http://enigma.ini.usc.edu/ongoing/enigma-anxiety/). All data access is contingent on approval by PIs from contributing samples.

## Acknowledgements

We would like to acknowledge the late Professor Dan J. Stein for his longstanding leadership and collaboration within ENIGMA-Anxiety Working Group and for his invaluable contributions to psychiatry, neuroscience, and mental health research. His intellectual generosity, mentorship, and unwavering support shaped this work and the broader field in lasting ways. His scientific legacy and his kindness to colleagues continue to inspire us.

ENIGMA extends gratitude to the NIH Big Data to Knowledge (BD2K) award for its foundational support and contributions to consortium development (U54 EB020403 awarded to PMT). Research reported in this publication was supported by NIH S10OD032285. For a comprehensive list of ENIGMA-related grant support, please visit: https://enigma.ini.usc.edu/about-2/funding/. SIT was supported by R01NS122827. KVB was supported by the Oppenheimer Memorial Trust (OMT Ref. 22148/01) and the University of Cape Town Postgraduate Publication Incentive. YBH and NY were funded by Joy Ventures (grant R01-30-2017-8156), and NY was partially funded by NIMH-Intramural Project, ZIAMH002781. KBB was supported by German Research Foundation (DFG), grant number: BE 3809/8-1, 604237. MC acknowledges support from the Ramón y Cajal program (RYC2024-050082-I) funded by MICIU/AEI/10.13039/501100011033 and by ESF+. CRKC was supported by R01MH129742, R01AG058854-05, R21MH139001, 5R01AG058854, research reported in this publication was supported by NIH S10OD032285. SHC was supported by MIST (grant number: RS-2023-00265883, RS-2025-16071556). UD was funded by the DFG (grant FOR2107 DA1151/5-1, DA1151/5-2, DA1151/9-1, DA1151/10-1, DA1151/11-1 to UD; SFB/TRR 393, project grant no 521379614) and the Interdisciplinary Center for Clinical Research (IZKF) of the medical faculty of Münster (grant Dan3/016/26 to UD). AGGD was financially supported by the South African Medical Research Council, Nuclear Technologies in Medicine and the Biosciences Initiative (NTeMBI) and the Harry Crossley Foundation. GAF was supported by R01MH132784, R01MH129694, R01MH125886, and a philanthropic gift from the Effie and Wofford Cain Foundation. BJH was supported by National Health and Medical Research Council (NHMRC) Project Grants 1064643 and 1145010. YH was supported by the AMED Brain/MINDS Beyond Program (Grant Number: JP18dm0307002), JSPS KAKENHI (Grant Numbers: 19K03309, 23K02956, 23K070004, 23K22361, 24K21493, 24K06547, 25K00879, 25K06842, 25H01085). HJ, AJ, TK, BS and FT were funded in part by the consortia grants from the German Research Foundation (DFG) FOR 2107, SFB/TRR 393 (“Trajectories of Affective Disorders”, project grant no 521379614), and Germany’s Excellence Strategy (EXC 3066/1 “The Adaptive Mind”, Project No. 533717223), as well as the DYNAMIC center, funded by the LOEWE program of the Hessian Ministry of Science and Arts (grant number: LOEWE1/16/519/03/09.001(0009)/98). EJL was funded by the DFG (SFB/TRR 393, project grant no 521379614) and the Medical faculty of Münster (grant LE121703). UL was funded in part by funded by the DFG – Projektnummer 44541416 – CRC TRR 58 “Fear, Anxiety, Anxiety Disorders” (project C09). KNTM was supported by StratNeuro, Karolinska Institutet, and the Swedish Research Council (Grant Nos. 2018-06729 and 2023-01362). SM was funded by the DFG, SFB/TRR393 (Project-ID 521379614) as well as ME62262 and the Innovative Medical Research (IMF) of the medical faculty of the University of Münster (grant no ME122205). IN was supported by grants of the DFG (grants NE 2254/1-2, NE 2254/2-1, NE 2254/3-1, NE 2254/4-1) and Forschungscampus Mittelhessen (FCMH). HO and RS: This study is in part the result of work supported with resources and the use of facilities at the Michael E. DeBakey Veterans Affairs Medical Center. These data included herein were collected through the use of facilities and resources at The Menninger Clinic, Houston, Texas USA. The content is solely the responsibility of the authors and does not represent the official views of the Department of Veterans Affairs or the United States government. RS was supported by the McNair Foundation (MIND_MB). VHA (I01CX001937). TMS: TSl was funded by the DFG (SFB/TRR 393, project grant no 521379614). RAV was funded by the Medical faculty of Münster (grant LE112403). XZ was supported by NIH K01MH122774, and NIH R01MH133840. DP and the associated work he performed was supported by Project ZIA-MH002782 in the National Institute of Mental Health Intramural Research Program. This research was supported [in part] by the Intramural Research Program of the National Institutes of Health (NIH). The contributions of the NIH author(s) are considered Works of the United States Government. The findings and conclusions presented in this paper are those of the author(s) and do not necessarily reflect the views of the NIH or the U.S. Department of Health and Human Services. MA was supported by The Netherlands Organization for Health Research and Development—ZonMw (research fellowship number 06360322210035), Dutch Research Council NWO (SSH Open Competition number 15810 & 20171), Leiden University Fund (project Youth Mental Health Meets Big Data Analytics, grant number LUF23075-5-306), Leiden University Fund (project grant number W213085-5). PMT and SIT were funded in part by NIH grants R01MH134962, R01MH131806, and R01MH138569. JMBH’s work is made possible by a grant from NeurolabNL (Small projects for NWA routes 21/22;NWA.1418.22.025) and past support from the Dutch Research Council NWO (Rubicon grant 019.201SG.022) and Medical Delta (NL; Talent Acceleration grant).

## References

1. American Psychiatric Association. (2013). Diagnostic and statistical manual of mental disorders (5th ed.). Washington, DC

2. Ruscio, A. M., Brown, T. A., Chiu, W. T., Sareen, J., Stein, M. B., & Kessler, R. C. (2008). Social fears and social phobia in the United States: Results from the National Comorbidity Survey Replication. Psychological Medicine, 38(1), 15–28. 10.1017/S0033291707001699

3. Stein, D. J., Lim, C. C. W., Roest, A. M., Jonge, P. De, Aguilar-gaxiola, S., Al-hamzawi, A., … Girolamo, G. De. (2017). The cross-national epidemiology of social anxiety disorder: Data from the World Mental Health Survey Initiative. BMC Medicine, 15(143), 1–21. 10.1186/s12916-017-0889-2

4. Stein, M. B., & Stein, D. J. (2008). Social anxiety disorder. The Lancet, 371(9618), 1115–1125. 10.1016/S0140-6736(08)60488-2 External Link

5. Rodriguez, B. F., Weisberg, R. B., Pagano, M. E., Machan, J. T., Culpepper, L., & Keller, M. B. (2004). Frequency and patterns of psychiatric comorbidity in a sample of primary care patients with anxiety disorders. Comprehensive Psychiatry, 45(2), 129–137. 10.1016/j.comppsych.2003.09.005

6. Meier, S. M., Mattheisen, M., Mors, O., Mortensen, P. B., Laursen, T. M., & Penninx, B. W. (2016). Increased mortality among people with anxiety disorders: Total population study. The British Journal of Psychiatry, 209(3), 216–221. 10.1192/bjp.bp.115.171975

7. Tully, P. J., & Cosh, S. M. (2023). Treating depression and anxiety disorders simultaneously in patients with cardiovascular diseases utilizing the unified protocol. Applications of the Unified Protocol in Health Conditions, 285.

8. Baur, V., Hänggi, J., Rufer, M., Delsignore, A., Jäncke, L., Herwig, U., & Brühl, A. B. (2011). White matter alterations in social anxiety disorder. Journal of Psychiatric Research, 45(10), 1366–1372. 10.1016/j.jpsychires.2011.05.007

9. Perna, G., Iannone, G., Alciati, A., & Caldirola, D. (2016). Are anxiety disorders associated with accelerated aging? A focus on neuroprogression. Neural Plasticity. 10.1155/2016/8457612

10. Bas-Hoogendam, J. M., Steenbergen, H. Van, Pannekoek, J. N., Fouche, J., Lochner, C., Hattingh, C. J., Cremers, H. R., Furmark, T., Månsson, K. N., Frick, A., Engman, J., Boraxbekk, C., Carlbring, P., Andersson, G., Fredrikson, M., Straube, T., Peterburs, J., Klumpp, H., Phan, K. L. … Van Der Wee, N. J. A. (2017). Voxel-based morphometry multi-center mega-analysis of brain structure in social anxiety disorder. NeuroImage: Clinical, 16, 678–688. 10.1016/j.nicl.2017.08.001

11. Groenewold, N. A., Bas-Hoogendam, J. M., Amod, A. R., Laansma, M. A., Van Velzen, L. S., Aghajani, M., … & Van der Wee, N. J. (2023). Volume of subcortical brain regions in social anxiety disorder: mega-analytic results from 37 samples in the ENIGMA-Anxiety Working Group. Molecular Psychiatry, 28(3), 1079–1089. 10.1038/s41380-022-01933-9

12. Irle, E., Ruhleder, M., Lange, C., Seidler-brandler, U., Salzer, S., Dechent, P., Weniger, G., Leibing, E., & Leichsenring, F. (2010). Reduced amygdalar and hippocampal size in adults with generalized social phobia. Journal of Psychiatry and Neuroscience, 35(2), 126–131. 10.1503/jpn.090041

13. Machado-de-Sousa, J. P., de Lima Osório, F., Jackowski, A. P., Bressan, R. A., Chagas, M. H., Torro-Alves, N., DePaula, A., Crippa, J. A. S. & Hallak, J. E. (2014). Increased amygdalar and hippocampal volumes in young adults with social anxiety. PloS one, 9(2). 10.1371/journal.pone.0088523

14. Zhao, Y., Chen, L., Zhang, W., Xiao, Y., Shah, C., Zhu, H., Yuan, M., Sun, H., Yue, Q., Jia, Z., Zhang, W., Kuang, W., Gong, Q., & Lui, S. (2017). Gray matter abnormalities in non-comorbid medication-naive patients with major depressive disorder or social anxiety disorder. EBioMedicine, 21, 228–235. 10.1016/j.ebiom.2017.06.013

15. Bas-Hoogendam, J. M., Groenewold, N. A., Aghajani, M., Freitag, G. F., Harrewijn, A., Hilbert, K., … & Suchan. (2020). ENIGMA-anxiety working group: Rationale for and organization of large-scale neuroimaging studies of anxiety disorders. Human Brain Mapping, 43(1), 83–112. 10.1002/hbm.25100

16. De Lange, A. M. G., Anatürk, M., Suri, S., Kaufmann, T., Cole, J. H., Griffanti, L., … & Ebmeier, K. P. (2020). Multimodal brain-age prediction and cardiovascular risk: The Whitehall II MRI sub-study. NeuroImage, 222, 117292. 10.1016/j.neuroimage.2020.117292

17. Franke, K., Ziegler, G., Klöppel, S., Gaser, C., & Alzheimer’s Disease Neuroimaging Initiative. (2010). Estimating the age of healthy subjects from T1-weighted MRI scans using kernel methods: Exploring the influence of various parameters. Neuroimage, 50(3), 883–892. 10.1016/j.neuroimage.2010.01.005

18. Franke, K., & Gaser, C. (2019). Ten years of BrainAGE as a neuroimaging biomarker of brain aging: What insights have we gained? Frontiers in Neurology, 10, 789. 10.3389/fneur.2019.00789

19. Han, L. K., Dinga, R., Hahn, T., Ching, C. R., Eyler, L. T., Aftanas, L., Aghajani, M., Aleman, A., Baune, B. T., Berger, K., Brak, I., Filho, G. B., Carballedo, A., Connolly, C. G., Couvy-Duchesne, B., Cullen, K. R., Dannlowski, U., Davey, C. G., & Dima, D. (2020). Brain aging in major depressive disorder: Results from the ENIGMA major depressive disorder working group. Molecular Psychiatry, 1–16. 10.1038/s41380-020-0754-0

20. Cole, J. H., & Franke, K. (2017). Predicting age using neuroimaging: Innovative brain ageing biomarkers. Trends in Neurosciences, 40(12), 681–690. 10.1016/j.tins.2017.10.001

21. Cole, J. H., Marioni, R. E., Harris, S. E., & Deary, I. J. (2018). Brain age and other bodily ‘ages’: Implications for neuropsychiatry. Molecular Psychiatry, 24(2), 266–281. 10.1038/s41380-018-0098-1

22. Ballester, P. L., Romano, M. T., de Azevedo Cardoso, T., Hassel, S., Strother, S. C., Kennedy, S. H., & Frey, B. N. (2022). Brain age in mood and psychotic disorders: a systematic review and meta-analysis. Acta Psychiatrica Scandinavica, 145(1), 42–55. 10.1111/acps.13371

23. Drobinin, V., Van Gestel, H., Helmick, C. A., Schmidt, M. H., Bowen, C. V., & Uher, R. (2022). The developmental brain age is associated with adversity, depression, and functional outcomes among adolescents. Biological Psychiatry: Cognitive Neuroscience and Neuroimaging, 7(4), 406–414. 10.1016/j.bpsc.2021.09.004

24. Han, L. K., Schnack, H. G., Brouwer, R. M., Veltman, D. J., van der Wee, N. J., van Tol, M. J., … & Penninx, B. W. (2021). Contributing factors to advanced brain aging in depression and anxiety disorders. Translational Psychiatry, 11(1), 1–11. 10.1038/s41398-021-01524-2

25. Kaufmann, T., van der Meer, D., et al (2019). Common brain disorders are associated with heritable patterns of apparent aging of the brain. Nature Neuroscience, 22(10), 1617–1623. 10.1038/s41593-019-0471-7

26. Lund, M. J., Alnæs, D., de Lange, A. M. G., Andreassen, O. A., Westlye, L. T., & Kaufmann, T. (2022). Brain age prediction using fMRI network coupling in youths and associations with psychiatric symptoms. NeuroImage: Clinical, 33, 102921. 10.1016/j.nicl.2021.102921

27. Zhou, Z., Li, H., Srinivasan, D., Abdulkadir, A., Nasrallah, I. M., Wen, J., … & ISTAGING Consortium. (2023). Multiscale functional connectivity patterns of the aging brain learned from harmonized rsfMRI data of the multi-cohort iSTAGING study. NeuroImage, 269, 119911. 10.1016/j.neuroimage.2023.119911

28. Koutsouleris, N., Davatzikos, C., Borgwardt, S., Gaser, C., Bottlender, R., Frodl, T., … & Meisenzahl, E. (2014). Accelerated brain aging in schizophrenia and beyond: A neuroanatomical marker of psychiatric disorders. Schizophrenia Bulletin, 40(5), 1140–1153. 10.1093/schbul/sbt142

29. Lieslehto, J., Jääskeläinen, E., Kiviniemi, V., Haapea, M., Jones, P. B., Murray, G. K., … & Koutsouleris, N. (2021).The progression of disorder-specific brain pattern expression in schizophrenia over 9 years. npj Schizophrenia, 7(1), 1–11. 10.1038/s41537-021-00157-0

30. McWhinney, S., Kolenic, M., Franke, K., Fialova, M., Knytl, P., Matejka, M., … & Hajek, T. (2021). Obesity as a risk factor for accelerated brain aging in first-episode psychosis—A longitudinal study. Schizophrenia Bulletin, 47(6), 1772–1781. 10.1093/schbul/sbab064

31. Van Gestel, H., Franke, K., Petite, J., Slaney, C., Garnham, J., Helmick, C., … & Hajek, T. (2019). Brain age in bipolar disorders: Effects of lithium treatment. Australian and New Zealand Journal of Psychiatry, 53(12), 1179–1188. 10.1177/0004867419857814

32. Blake, K. V., Hilbert, K., Ipser, J. C., Han, L. K., Bas-Hoogendam, J. M., Åhs, F., … & Groenewold, N. A. (2025). Brain aging in specific phobia: An ENIGMA-anxiety mega-analysis. MedRxiv, 2025-03. 10.1101/2025.03.19.25323474

33. Richier, C., Zugman, A., Harrewijn, A., Cardinale, E. M., Khosravi, P., Aghajani, M., … & Sawyers, C. K. (2026). Brain age prediction in generalized anxiety disorder using a convolutional neural network. Translational Psychiatry. 10.1038/s41398-026-04078-3

34. Fischl, B., Salat, D. H., Busa, E., Albert, M., Dieterich, M., Haselgrove, C., … & Dale, A. M. (2002). Whole brain segmentation: Automated labeling of neuroanatomical structures in the human brain. Neuron, 33(3), 341–355. 10.1016/S0896-6273(02)00569-X

35. Grasby, K. L., Jahanshad, N., Painter, J. N., Colodro-Conde, L., Bralten, J., Hibar, D. P., … & Genetics working group. (2020). The genetic architecture of the human cerebral cortex. Science, 367(6484). 10.1126/science.aay6690

36. Stein, J. L., Medland, S. E., Vasquez, A. A., Hibar, D. P., Senstad, R. E., Winkler, A. M., Toro, R., Appel, K., Bartecek, R., Bergman, O., Bernard, M., Brown, A. A., Cannon, D. M., Chakravarty, M. M., Christoforou, A., Domin, M., Grimm, O., Hollinshead, M., Holmes, A. J. …Thompson, P. M. (2012). Identification of common variants associated with human hippocampal and intracranial volumes. Nature Genetics, 44(5), 552–561. 10.1038/ng.2250

37. Pedregosa, F., Varoquaux, G., Gramfort, A., Michel, V., Thirion, B., Grisel, O., … & Duchesnay, E. (2011). Scikit-learn: Machine learning in Python. the Journal of machine Learning research, 12, 2825–2830.

38. Desikan, R. S., Ségonne, F., Fischl, B., Quinn, B. T., Dickerson, B. C., Blacker, D., … & Killiany, R. J. (2006). An automated labeling system for subdividing the human cerebral cortex on MRI scans into gyral based regions of interest. Neuroimage, 31(3), 968–980. 10.1016/j.neuroimage.2006.01.021

39. Boedhoe, P. S., Schmaal, L., Abe, Y., Ameis, S. H., Arnold, P. D., Batistuzzo, M. C., … & members of the ENIGMA OCD Working Group. (2017). Distinct subcortical volume alterations in pediatric and adult OCD: A worldwide meta-and mega-analysis. American Journal of Psychiatry, 174(1), 60–69. 10.1176/appi.ajp.2016.16020201

40. Boedhoe, P. S., Heymans, M. W., Schmaal, L., Abe, Y., Alonso, P., Ameis, S. H., … & Twisk, J. W. (2019). An empirical comparison of meta-and mega-analysis with data from the ENIGMA obsessive-compulsive disorder working group. Frontiers in Neuroinformatics, 12, 102. 10.3389/fninf.2018.00102.

41. Zugman, A., Harrewijn, A., Cardinale, E. M., Zwiebel, H., Freitag, G. F., Werwath, K. E., … & Winkler, A. M. (2022). Mega-analysis methods in ENIGMA: The experience of the generalized anxiety disorder working group. Human Brain Mapping, 43(1). 10.1002/hbm.25096

42. Thoemmes, F. J., & Kim, E. S. (2011). A systematic review of propensity score methods in the social sciences. Multivariate Behavioral Research, 46(1), 90–118. 10.1080/00273171.2011.540475

43. Liebowitz, M. R. (1987). Social phobia. Modern problems of pharmacopsychiatry, 22(141), e173.

44. Spielberger, C. D., & Vagg, P. R. (1984). Psychometric properties of the STAI: A reply to Ramanaiah, Franzen, and Schill. Journal of Personality Assessment, 48(1), 95–97. 10.1207/s15327752jpa4801_16

45. Constantinides, C., Han, L. K., Alloza, C., Antonucci, L. A., Arango, C., Ayesa-Arriola, R., … & Walton, E. (2023). Brain ageing in schizophrenia: Evidence from 26 international cohorts via the ENIGMA Schizophrenia consortium. Molecular Psychiatry, 28(3), 1201–1209. 10.1038/s41380-022-01897-w

46. Butler, E. R., Chen, A., Ramadan, R., Le, T. T., Ruparel, K., Moore, T. M., … & Shinohara, R. T. (2021). Pitfalls in Brain Age Analyses (Vol. 42, No. 13, pp. 4092-4101). Hoboken, USA: John Wiley & Sons, Inc. 10.1002/hbm.25533

47. De Lange, A. M. G., & Cole, J. H. (2020). Commentary: Correction procedures in brain-age prediction. NeuroImage: Clinical, 26. 10.1016/j.nicl.2020.102229

48. De Lange, A. M. G., Anatürk, M., Rokicki, J., Han, L. K., Franke, K., Alnæs, D., … & Cole, J. H. (2022). Mind the gap: Performance metric evaluation in brain-age prediction. Human Brain Mapping, 43(10), 3113–3129. 10.1002/hbm.25837

49. De Vries, Y. A., Al-Hamzawi, A., Alonso, J., Borges, G., Bruffaerts, R., Bunting, B., … & De Jonge, P. (2019). Childhood generalized specific phobia as an early marker of internalizing psychopathology across the lifespan: Results from the World Mental Health Surveys. BMC Medicine, 17(1), 101. 10.1186/s12916-019-1328-3.

50. Wardenaar, K. J., Lim, C. C., Al-Hamzawi, A. O., Alonso, J., Andrade, L. H., Benjet, C. D., … & De Jonge, P. (2017). The cross-national epidemiology of specific phobia in the World Mental Health Surveys. Psychological Medicine, 47(10), 1744–1760. 10.1017/S0033291717000174.

51. Blake, K. V., Ntwatwa, Z., Kaufmann, T., Stein, D. J., Ipser, J. C., & Groenewold, N. A. (2023). Advanced brain aging in adult psychopathology: A systematic review and meta-analysis of structural MRI studies. Journal of Psychiatric Research. 10.1016/j.jpsychires.2022.11.011

52. Fedoce, A. D. G., Ferreira, F., Bota, R. G., Bonet-Costa, V., Sun, P. Y., & Davies, K. J. (2018). The role of oxidative stress in anxiety disorder: cause or consequence?. Free radical research, 52(7), 737–750. 10.1080/10715762.2018.1475733

53. Bouayed, J., Rammal, H., & Soulimani, R. (2009). Oxidative stress and anxiety: relationship and cellular pathways. Oxidative Medicine Cellular Longevity, 2, 63–67. 10.4161/oxim.2.2.7944

54. Tsaluchidu, S., Cocchi, M., Tonello, L., & Puri, B. K. (2008). Fatty acids and oxidative stress in psychiatric disorders. BMC Psychiatry, 8, 1–3. 10.1186/1471-244X-8-S1-S5

55. Peirce, J. M., & Alviña, K. (2019). The role of inflammation and the gut microbiome in depression and anxiety. Journal of Neuroscience Research, 97(10), 1223–1241. 10.1002/jnr.24476

56. Sherbourne, C. D., Sullivan, G., Craske, M. G., Roy-Byrne, P., Golinelli, D., Rose, R. D., … & Stein, M. B. (2010). Functioning and disability levels in primary care out-patients with one or more anxiety disorders. Psychological Medicine, 40(12), 2059–2068. 10.1017/S0033291710000176

57. Koyuncu, A., İnce, E., Ertekin, E., & Tükel, R. (2019). Comorbidity in social anxiety disorder: diagnostic and therapeutic challenges. Drugs in Context, 8, 212573. 10.7573/dic.212573

58. Miyazaki, M., Yoshino, A., & Nomura, S. (2011). Relationships between anxiety severity, diagnosis of multiple anxiety disorders, and comorbid major depressive disorder. Asian Journal of Psychiatry, 4(4), 293–296. 10.1016/j.ajp.2011.10.003

59. Hendriks, S. M., Spijker, J., Licht, C. M., Hardeveld, F., de Graaf, R., Batelaan, N. M., … & Beekman, A. T. (2016). Long-term disability in anxiety disorders. Bmc Psychiatry, 16, 1–8. 10.1186/s12888-016-0946-y

60. Rytwinski, N. K., Fresco, D. M., Heimberg, R. G., Coles, M. E., Liebowitz, M. R., Cissell, S., … & Hofmann, S. G. (2009). Screening for social anxiety disorder with the self-report version of the Liebowitz Social Anxiety Scale. Depression and anxiety, 26(1), 34–38. 10.1002/da.20503

61. Riley, R. D., Lambert, P. C., & Abo-Zaid, G. (2010). Meta-analysis of individual participant data: Rationale, conduct, and reporting. Bmj, 340. 10.1136/bmj.c221

62. Shahar, E., & Shahar, D. J. (2013). Causal diagrams and the cross-sectional study. Clinical Epidemiology, 57–65. 10.2147/CLEP.S42843

63. Wang, X., & Cheng, Z. (2020). Cross-sectional studies: strengths, weaknesses, and recommendations. Chest, 158(1), S65–S71. 10.1016/j.chest.2020.03.012

64. Jawinski, P., Markett, S., Drewelies, J., Düzel, S., Demuth, I., Steinhagen-Thiessen, E., … & Kühn, S. (2022). Linking brain age gap to mental and physical health in the berlin aging study ii. Frontiers in Aging Neuroscience, 14, 791222. 10.3389/fnagi.2022.791222

65. Tanveer, M., Ganaie, M. A., Beheshti, I., Goel, T., Ahmad, N., Lai, K. T., … & Lin, C. T. (2023). Deep learning for brain age estimation: A systematic review. Information Fusion, 96, 130–143. 10.1016/j.inffus.2023.03.007

