## Supplementary Materials for "Brain Ageing in Social Anxiety Disorder: An ENIGMA-Anxiety Mega-Analysis Across 26 International Cohorts"

**List of Tables and Figures**

Figure S1. Age density plot per diagnostic group.

Figure S2a-S2b. Age boxplots.

Figure S3a-S3b. Brain-PAD boxplots.

Figure S4a-S4b. MAE boxplots.

Figure S5a-S5b. Cortical thickness boxplots.

Table S1. Model fit statistics: by full sample, by diagnostic group, and by sex.

Table S2. Mean absolute error per study site and diagnostic group

Table S3. Pearson’s R and R^2^: brain age and age and brain-PAD and age, per study site and diagnostic group.

Figure S6. Brain age by age scatterplots per diagnostic group and study site.

Figure S7. Brain-PAD residuals plot for primary linear mixed-effects model without interaction term.

Table S4. Brain-PAD extreme values.

Figure S8. Boxplot of brain-PAD extreme values.

Table S5. Between-group differences (SAD vs HC) in brain-PAD after removal of n=2 outliers for brain-PAD.

Table S6. Between-group differences (SAD vs HC) in brain-PAD, models including interaction variables.

Table S7. Summary of balance for matched and unmatched data per study site.

Figure S9. Matched age density plots.

Table S9. Between-group differences (SAD vs HC) in brain-PAD, matched dataset.

Table S10. Sociodemographic information for SAD participants included in SAD (ANX comorbidity) sub-analysis.

Table S11. Sample overlap between SAD participant subgroups based on comorbidity and medication use.

Table S12. Between-group differences in brain-PAD, SAD with ANX comorbidity (n=105) vs HCs (n=1298), exclusion of NESDA sites.

Table S13. Between group differences in brain-PAD: SAD with ANX comorbidities, removal of specific diagnoses.

**Age Density Plot**

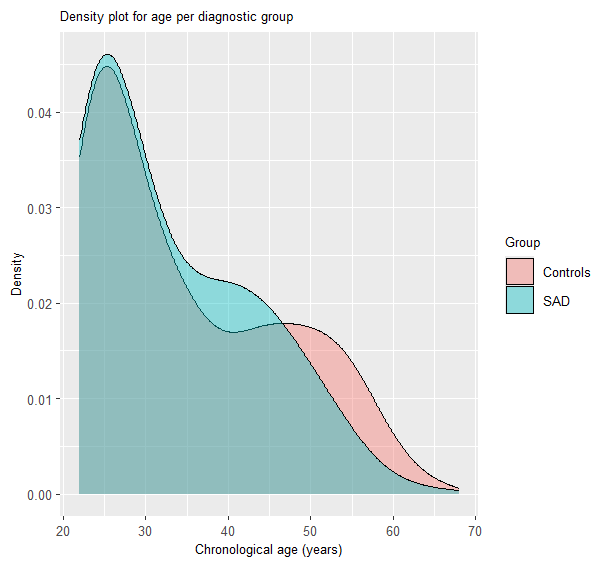

Figure S1. Age density plot per diagnostic group (SAD n=576, HC n=1,355). Note. HC, healthy controls; SAD, social anxiety disorder.

**Variable Distributions**

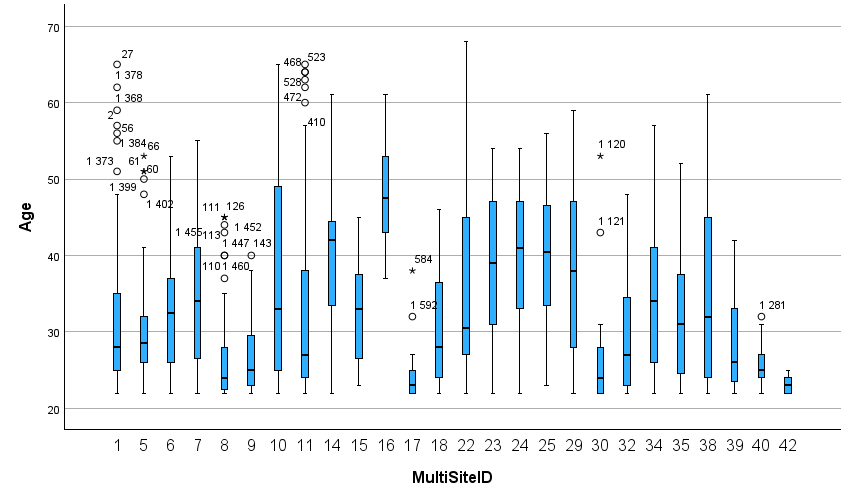

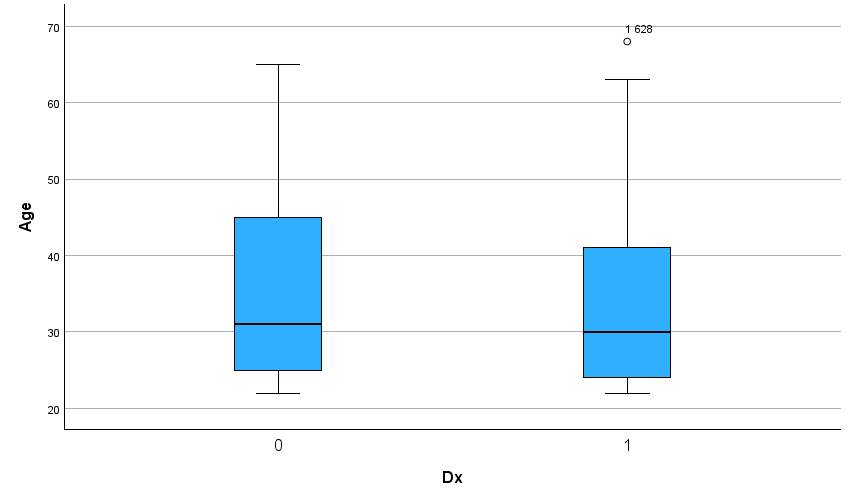
**Age Boxplots**

Figure S2a. Age boxplots per diagnostic group. Figure S2b. Age boxplots per research site. Note. Dx, diagnosis. Healthy controls = 0; Social anxiety disorder = 1. Circles and stars represent outliers (>1.5 x IQR beyond the quartiles) and extreme outliers (>3 x IQR beyond the quartiles), respectively.

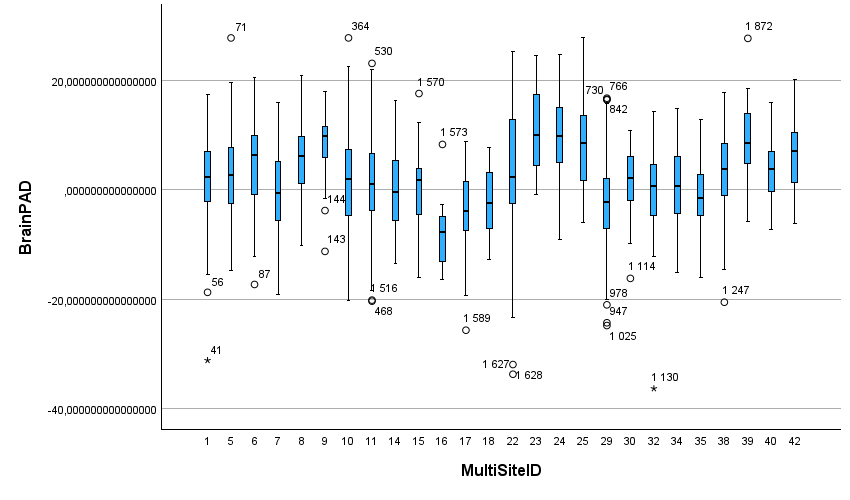

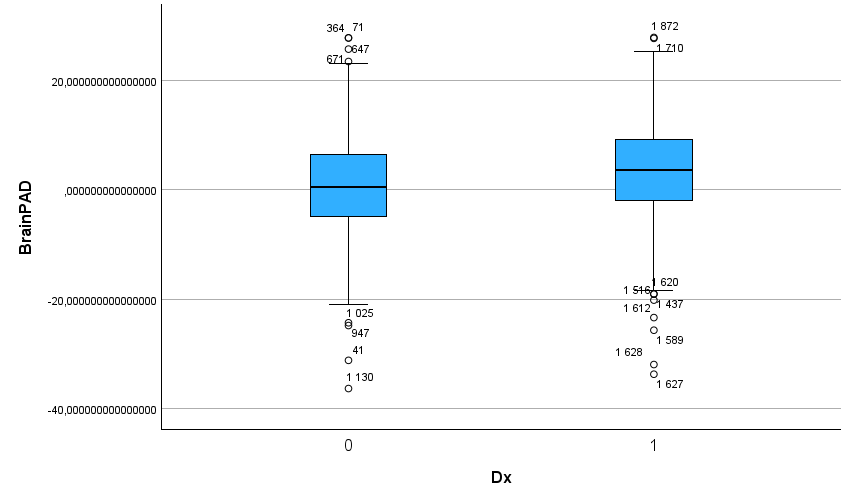
**Brain-PAD Boxplots**

Figure S3a. Brain-PAD boxplots per diagnostic group. Figure S3b. Brain-PAD boxplots per research site. Note. BrainPAD, brain predicted age difference; Dx, diagnosis. Healthy controls = 0; Social anxiety disorder = 1. Circles and stars represent outliers (>1.5 x IQR beyond the quartiles) and extreme outliers (>3 x IQR beyond the quartiles), respectively.

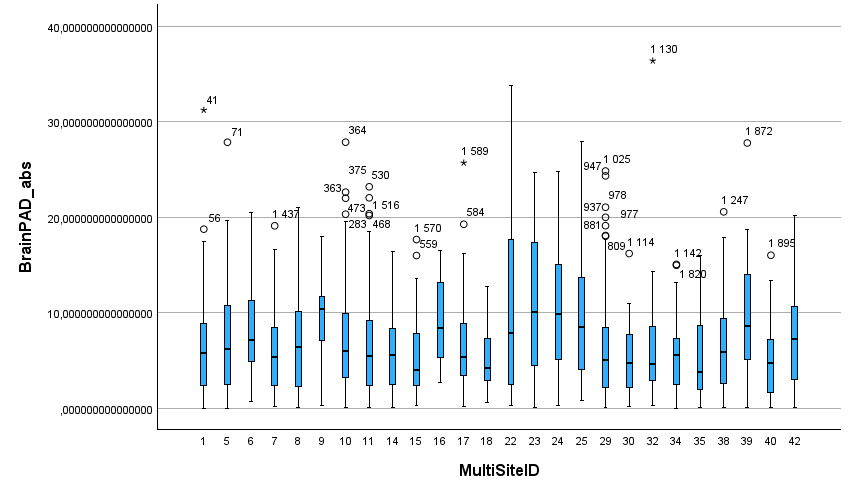

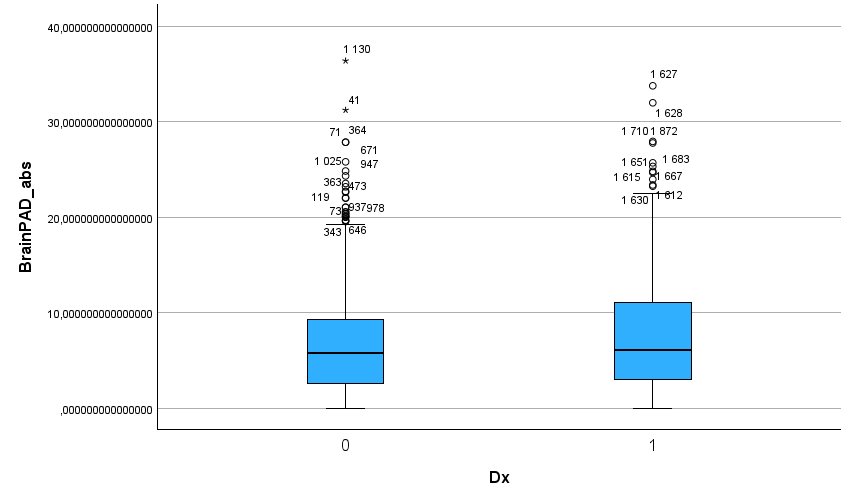
**MAE Boxplots**

Figure S4a. MAE boxplots per diagnostic group. Figure S4b. MAE boxplots per research site. Note. BrainPAD abs, brain predicted age difference absolute value/mean absolute error; Dx, diagnosis; MAE, mean absolute error. Healthy controls = 0; Social anxiety disorder = 1. Circles and stars represent outliers (>1.5 x IQR beyond the quartiles) and extreme outliers (>3 x IQR beyond the quartiles), respectively.

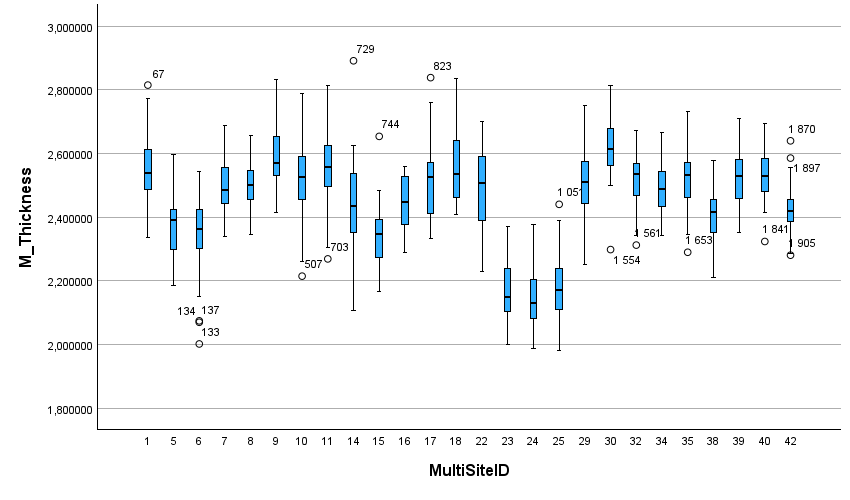

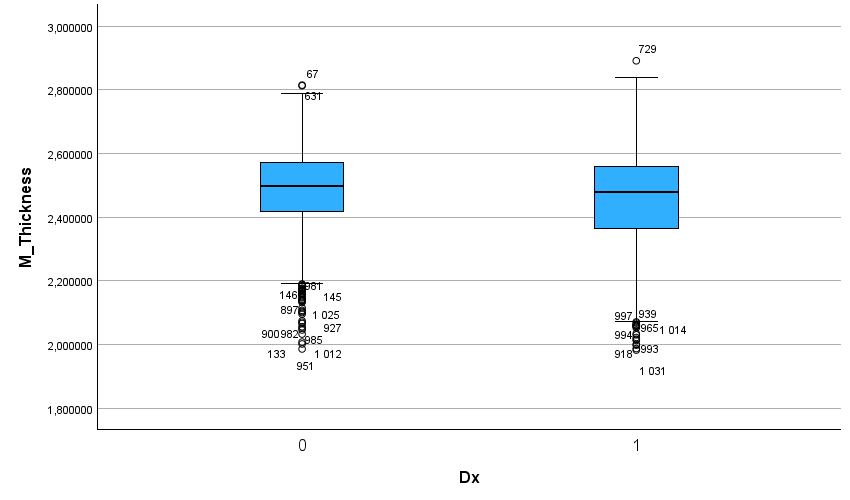
**Cortical Thickness Boxplots**

Figure S5a. Cortical thickness boxplots per diagnostic group. Figure S5b. Cortical thickness boxplots per research site. Note. Dx, diagnosis; M Thickness, mean cortical thickness. Healthy controls = 0; Social anxiety disorder = 1. Circles and stars represent outliers (>1.5 x IQR beyond the quartiles) and extreme outliers (>3 x IQR beyond the quartiles), respectively.

**Model Fit Statistics**

**MAE and Correlations, Full Sample and Per Diagnostic Group and Sex**

Table S1. Model fit statistics: by full sample, by diagnostic group, and by sex.

|  | Full sample, n=1,931 | SAD, n=576 | HC, n=1,355 | Males, n=779 | Females, n=1,152 |
| --- | --- | --- | --- | --- | --- |
| MAE (SD) | 6.90 (5.32) | 7.53 (5.81) | 6.62 (5.07) | 6.46 (5.02) | 7.19 (5.49) |
| R (brain age, age) | 0.69 | 0.65 | 0.72 | 0.71 | 0.68 |
| R^2^ (brain age, age) | 0.48 | 0.42 | 0.51 | 0.51 | 0.46 |
| R (brain-PAD, age) | 0.44 | 0.32 | 0.48 | 0.42 | 0.45 |
| R^2^ (brain-PAD, age) | 0.19 | 0.10 | 0.23 | 0.18 | 0.20 |

Note: brain-PAD, brain predicted age difference; HC, healthy control; MAE, mean absolute error; R, Pearson correlation; SAD, social anxiety disorder; SD, standard deviation.

**MAE Per Site and Diagnostic Group**

Table S2. Mean absolute error per study site and diagnostic group.

|  |  | HC | | SAD | |
| --- | --- | --- | --- | --- | --- |
| MultiSiteID | Site | **n** | **MAE (SD)** | **n** | **MAE (SD)** |
| 1 | BCM/MIND-MB | 59 | 5.84 (5.40) | 42 | 7.16 (4.93) |
| 38 | Chiba | 72 | 6.65 (4.60) | 15 | 6.61 (6.35) |
| 39 | Columbia MRT | 18 | 7.52 (5.05) | 26 | 11.15 (6.08) |
| 5 | Columbia SAD | 15 | 8.73 (7.61) | 15 | 5.91 (4.44) |
| 6 | Columbia SPP | 18 | 9.44 (5.61) | 16 | 7.35 (4.37) |
| 7 | DCCN | 17 | 5.71 (4.51) | 15 | 7.45 (5.82) |
| 8 | DelMar | 27 | 6.73 (5.09) | 24 | 7.15 (5.41) |
| 9 | Dresden | 19 | 9.37 (4.44) | 12 | 9.10 (4.87) |
| 10 | FOR2107 MR | 239 | 7.04 (5.02) | 24 | 6.68 (3.02) |
| 11 | FOR2107 MS | 141 | 6.12 (4.80) | 23 | 7.63 (5.75) |
| 14 | Houston | 11 | 4.84 (3.88) | 20 | 6.28 (3.83) |
| 15 | Istanbul | 20 | 5.70 (4.17) | 24 | 5.76 (4.54) |
| 16 | LFLSAD | 10 | 9.44 (4.02) | 10 | 8.74 (4.67) |
| 17 | UC Louvain | 13 | 7.37 (5.77) | 16 | 7.24 (6.63) |
| 18 | LUMC | 17 | 5.49 (3.83) | 15 | 4.70 (2.14) |
| 22 | MSAD | 17 | 9.24 (8.05) | 17 | 13.14 (10.82) |
| 23 | NESDA Amsterdam | 20 | 8.59 (6.03) | 34 | 12.33 (7.32) |
| 24 | NESDA Leiden | 26 | 10.26 (6.38) | 31 | 11.13 (6.20) |
| 25 | NESDA Groningen | 11 | 8.67 (7.19) | 33 | 10.12 (6.79) |
| 29 | SP Muenster | 430 | 5.99 (4.60) | 59 | 5.37 (4.31) |
| 30 | TIP | 11 | 5.30 (4.79) | 9 | 5.37 (3.77) |
| 32 | UCSD Sapient Insula | 20 | 6.72 (7.65) | 16 | 5.43 (3.88) |
| 34 | Umea Sofie | 20 | 6.44 (3.80) | 21 | 4.46 (3.77) |
| 35 | Umea Vox | 20 | 5.96 (5.52) | 19 | 4.86 (3.44) |
| 40 | Seoul SAD AC | 32 | 4.31 (3.47) | 28 | 5.83 (4.15) |
| 42 | YSAD | 52 | 8.03 (5.29) | 12 | 4.58 (3.61) |

Note. HC, healthy controls; MAE, mean absolute error; N, number; NA, not applicable; NS, not significant; SAD, social anxiety disorder; SD, standard deviation.

**Correlations Per Site and Diagnostic Group**

Table S3. Pearson’s R and R^2^: brain age and age and brain-PAD and age, per study site and diagnostic group.

|  |  | HC | |  | SAD | |  |
| --- | --- | --- | --- | --- | --- | --- | --- |
| Site | **Correlations** | **N** | **Brain age, age** | **Brain-PAD, age** | **N** | **Brain age, age** | **Brain-PAD, age** |
| BCM/MIND-MB | R | 59 | 0.58 | 0.57 | 42 | 0.72 | 0.69 |
|  | R^2^ |  | 0.33 | 0.33 |  | 0.52 | 0.41 |
| Chiba | R | 72 | 0.81 | 0.67 | 15 | 0.60 | 0.61 |
|  | R^2^ |  | 0.65 | 0.45 |  | 0.36 | 0.37 |
| Columbia MRT | R | 18 | 0.62 | 0.11 | 26 | 0.51 | 0.46 |
|  | R^2^ |  | 0.38 | 0.01 |  | 0.26 | 0.21 |
| Columbia SAD | R | 15 | 0.42 | 0.64 | 15 | 0.66 | 0.33 |
|  | R^2^ |  | 0.18 | 0.41 |  | 0.44 | 0.11 |
| Columbia SPP | R | 18 | 0.72 | 0.09 | 16 | 0.49 | 0.56 |
|  | R^2^ |  | 0.52 | 0.01 |  | 0.24 | 0.31 |
| DCCN | R | 17 | 0.73 | 0.54 | 15 | 0.51 | 0.47 |
|  | R^2^ |  | 0.53 | 0.29 |  | 0.26 | 0.22 |
| DelMar, | R | 27 | 0.68 | 0.23 | 24 | 0.64 | 0.38 |
|  | R^2^ |  | 0.46 | 0.05 |  | 0.41 | 0.15 |
| Dresden | R | 19 | 0.07 | 0.73 | 12 | 0.71 | 0.01 |
|  | R^2^ |  | 0.01 | 0.54 |  | 0.50 | <0.00 |
| FOR2107 MR | R | 239 | 0.74 | 0.58 | 24 | 0.72 | 0.28 |
|  | R^2^ |  | 0.55 | 0.34 |  | 0.52 | 0.08 |
| FOR2107 MS | R | 141 | 0.75 | 0.49 | 23 | 0.70 | 0.48 |
|  | R^2^ |  | 0.56 | 0.24 |  | 0.49 | 0.24 |
| Houston | R | 11 | 0.69 | 0.49 | 20 | 0.73 | 0.66 |
|  | R^2^ |  | 0.47 | 0.24 |  | 0.53 | 0.44 |
| Istanbul | R | 20 | 0.64 | 0.02 | 24 | 0.74 | 0.16 |
|  | R^2^ |  | 0.41 | <0.00 |  | 0.55 | 0.03 |
| LFLSAD | R | 10 | 0.85 | 0.47 | 10 | 0.30 | 0.35 |
|  | R^2^ |  | 0.73 | 0.22 |  | 0.09 | 0.21 |
| UC Louvain | R | 13 | 0.17 | 0.61 | 16 | 0.30 | 0.01 |
|  | R^2^ |  | 0.03 | 0.38 |  | 0.09 | <0.00 |
| LUMC | R | 17 | 0.77 | 0.07 | 15 | 0.77 | 0.19 |
|  | R^2^ |  | 0.60 | 0.01 |  | 0.58 | 0.44 |
| MSAD | R | 17 | 0.20 | 0.60 | 17 | 0.16 | 0.72 |
|  | R^2^ |  | 0.04 | 0.36 |  | 0.03 | 0.52 |
| NESDA Amsterdam | R | 20 | 0.79 | 0.32 | 34 | 0.74 | 0.20 |
|  | R^2^ |  | 0.62 | 0.10 |  | 0.55 | 0.04 |
| NESDA Leiden | R | 26 | 0.70 | 0.39 | 31 | 0.81 | 0.01 |
|  | R^2^ |  | 0.43 | 0.16 |  | 0.65 | <0.00 |
| NESDA Groningen | R | 11 | 0.51 | 0.58 | 33 | 0.60 | 0.44 |
|  | R^2^ |  | 0.26 | 0.34 |  | 0.36 | 0.20 |
| SP Muenster | R | 430 | 0.78 | 0.48 | 59 | 0.76 | 0.41 |
|  | R^2^ |  | 0.61 | 0.23 |  | 0.58 | 0.17 |
| TIP | R | 11 | 0.75 | 0.37 | 9 | 0.90 | 0.78 |
|  | R^2^ |  | 0.56 | 0.14 |  | 0.82 | 0.61 |
| UCSD Sapient Insula | R | 20 | 0.43 | 0.31 | 16 | 0.66 | 0.61 |
|  | R^2^ |  | 0.19 | 0.10 |  | 0.43 | 0.38 |
| Umea Sofie | R | 20 | 0.71 | 0.39 | 21 | 0.75 | 0.67 |
|  | R^2^ |  | 0.51 | 0.15 |  | 0.56 | 0.45 |
| Umea Vox | R | 20 | 0.75 | 0.62 | 19 | 0.60 | 0.44 |
|  | R^2^ |  | 0.56 | 0.38 |  | 0.36 | 0.20 |
| Seoul SAD AC | R | 32 | 0.38 | 0.09 | 28 | 0.27 | 0.21 |
|  | R^2^ |  | 0.14 | 0.01 |  | 0.07 | 0.05 |
| YSAD | R | 52 | 0.18 | 0.01 | 12 | 0.41 | 0.23 |
|  | R^2^ |  | 0.03 | <0.00 |  | 0.17 | 0.05 |

Note. brain-PAD, brain predicted age difference; HC, healthy controls; N, number; NA, not applicable; R, Pearson’s R; R^2^, Pearson’s R-squared; SAD, social anxiety disorder.

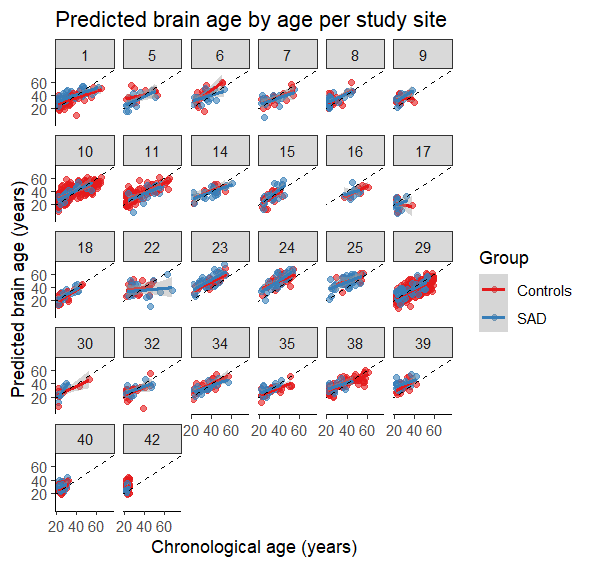
**Brain Age by Age Scatterplots**

Figure S6. Brain age by age scatterplots per diagnostic group and study site.

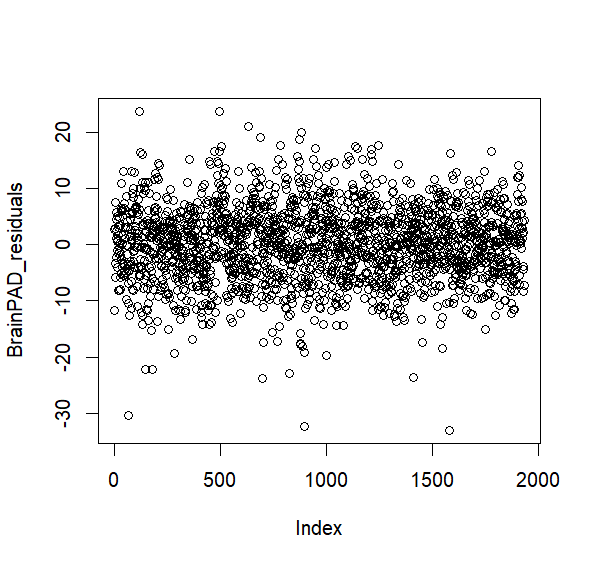
**Brain-PAD Residuals Plot**

Figure S7. Brain-PAD residuals plot for primary linear mixed-effects model without interaction term.

**Brain-PAD Extreme Values**

The below table and boxplot demonstrate the highest and lowest extreme brain-PAD values, generated with SPSS.

Table S4. Brain-PAD extreme values.

|  | Case number | Site | Brain-PAD value |
| --- | --- | --- | --- |
| Highest | 1710 | NESDA Groningen | 27.94 |
| Lowest | 1130 | UCSD Sapient Insula | -36.38 |

Note. Brain-PAD, brain predicted age difference.

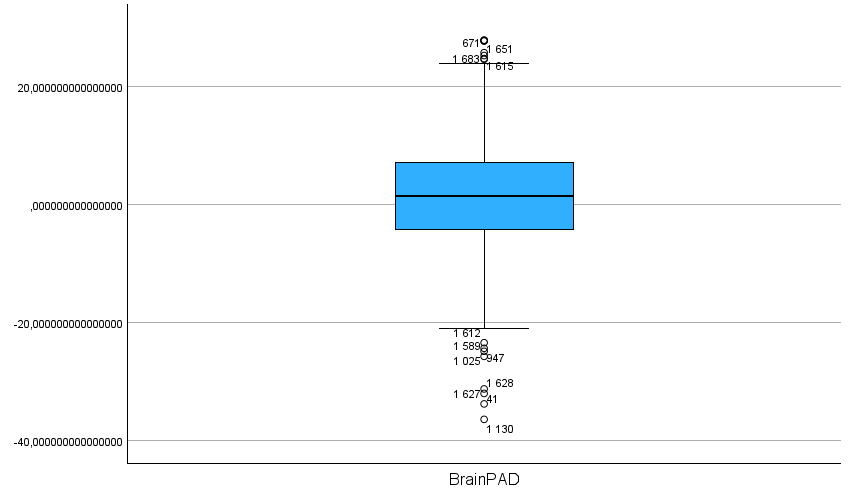

Figure S8. Boxplot of brain-PAD extreme values. Note. BrainPAD, brain predicted age difference. Circles and stars represent outliers (>1.5 x IQR beyond the quartiles) and extreme outliers (>3 x IQR beyond the quartiles), respectively.

**Linear Mixed-Effects Model: Brain-PAD Between Groups with Outlier Removal (1 SAD; 1 HC)**

Table S5. Between-group differences (SAD vs HC) in brain-PAD after removal of n=2 outliers for brain-PAD.

|  | 𝛃 (SE) | t-value | *p* |
| --- | --- | --- | --- |
| Intercept (site) | 2.32(0.95) | 2.44 | **0.015** |
| Diagnosis | 0.64(0.37) | 1.73 | 0.084 |
| Sex | -0.15(0.32) | -0.48 | 0.634 |
| AgeC | -0.32(0.02) | -16.10 | **<0.001** |
| AgeC^2^ | -4.68 x 10^-3^ (1.46 x 10^-3^) | -3.20 | **0.001** |

Note: AgeC, mean-centred chronological age; AgeC^2^, mean-centred chronological age squared; HC, healthy controls; CI, confidence intervals; SAD, social anxiety disorder; SE, standard error. 𝛃 is measured in years.

**Linear Mixed-Effects Model: Brain-PAD Between Groups with Interaction Variables**

Table S6. Between-group differences (SAD vs HC) in brain-PAD, models including interaction variables.

|  | Dx | | AgeC-by-dx | | AgeC^2^-by-Dx | | Sex-by-dx | |
| --- | --- | --- | --- | --- | --- | --- | --- | --- |
| Model | **𝛃 (SE)** | ***p*** | **𝛃 (SE)** | ***p*** | **𝛃 (SE)** | ***p*** | **𝛃 (SE)** | ***p*** |
| Fit 1b | 1.22(0.53) | **0.021** | -0.01(0.04) | 0.865 | -0.01(0.00) | 0.129 |  |  |
| Fit 1c | 1.53(0.67) | **0.020** | -0.01(0.04) | 0.806 | -0.01(0.00) | 0.147 | -0.55(0.70) | 0.428 |
| Fit 1d | 0.65(0.37) | 0.081 | -0.034(0.03) | 0.282 |  |  |  |  |
| Fit 1e | 1.26(0.48) | **0.009** |  |  | -0.01(0.00) | 0.064 |  |  |
| Fit 1f | 1.05(0.56) | 0.062 |  |  |  |  | -0.58(0.69) | 0.404 |

Note. Dx, diagnosis; AgeC-by-dx, mean-centred chronological age by diagnosis interaction term; AgeC^2^-by-dx, mean-centred chronological age squared by diagnosis interaction term; SE, standard error; Sex-by-dx, sex by diagnosis interaction term. 𝛃 is measured in years. Each model fit represents linear mixed effects models with different interaction variables and combinations included.

**Primary Linear Mixed-Effects Model Results in Current SAD (Not Lifetime) and HCs**

Table S7. Between-group differences in brain-PAD, current SAD (n=561) vs HCs (n=1,355)

|  | 𝛃 (SE) | t-value | *p* | Cohen’s *d*, (95% CI) |
| --- | --- | --- | --- | --- |
| Intercept (site) | 2.26(0.98) | 2.32 | **0.020** |  |
| Diagnosis | 0.72(0.38) | 1.92 | 0.056 | 0.08(-0.01-0.19) |
| Sex | -0.17(0.32) | -0.53 | 0.595 |  |
| AgeC | -0.32(0.02) | -16.14 | **<0.001** |  |
| AgeC^2^ | -0.01(0.00) | -2.92 | **0.004** |  |

Note: AgeC, mean-centred chronological age; AgeC^2^, mean-centred chronological age squared; HC, healthy controls; CI, confidence intervals; SAD, social anxiety disorder; SE, standard error. 𝛃 is measured in years.

**Propensity Score Matching**

**Matching Methods and Summary of Balance**

Table S8. Summary of balance for matched and unmatched data per study site.

| Study site | Variable | SAD means | HC means | Std. mean difference | Variance ratios | eCDF mean | eCDF maximum | Pair distances |
| --- | --- | --- | --- | --- | --- | --- | --- | --- |
| FOR2107 MS unmatched *(SAD n=23; HC n=141)* | **Distance** | 0.23 | 0.13 | 0.84 | 1.10 | 0.27 | 0.51 | NA |
|  | **Age** | 42.04 | 30.97 | 0.96 | 1.11 | 0.27 | 0.51 | NA |
| FOR2107 MS matched *(SAD n=23; HC n=37)* | **Distance** | 0.23 | 0.22 | 0.07 | 1.05 | 0.02 | 0.11 | 0.09 |
|  | **Age** | 42.04 | 41.35 | 0.06 | 1.05 | 0.02 | 0.11 | 0.07 |
| SP Muenster unmatched *(SAD n=59; HC n=430)* | **Distance** | 0.12 | 0.12 | 0.44 | 0.96 | 0.14 | 0.24 | NA |
|  | **Age** | 34.07 | 38.79 | -0.47 | 0.84 | 0.14 | 0.24 | NA |
| SP Muenster matched *(SAD n=59; HC n=118)* | **Distance** | 0.14 | 0.14 | <0.00 | 1.01 | <0.00 | 0.01 | <0.00 |
|  | **Age** | 34.07 | 34.08 | <-0.00 | 1.01 | <0.00 | 0.01 | <0.00 |

Note. eCDF, empirical cumulative distribution function statistics; HC, healthy control; SAD, social anxiety disorder; std, standardised.

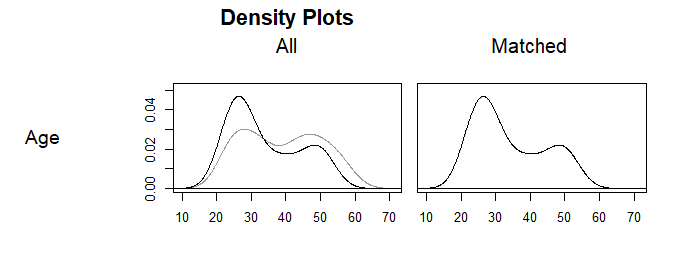

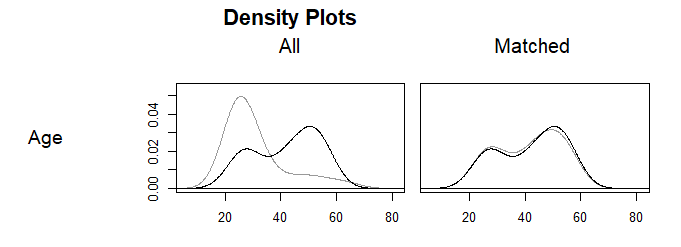
**FOR2107MS	SP Muenster**

Figure S9. Matched age density plots. Grey lines represent the control group; black lines represent the SAD group.

**Primary Linear Mixed-Effects Model Results in Matched Dataset**

Table S9. Between-group differences (SAD vs HC) in brain-PAD, propensity score matched dataset.

|  | 𝛃 (SE) | t-value | *p* | Cohen’s *d* |
| --- | --- | --- | --- | --- |
| Intercept (site) | 2.46(0.97) | 2.52 | **0.012** |  |
| Diagnosis | 0.69(3.90) | 1.78 | 0.075 | 0.08(-0.02-0.18) |
| Sex | 0.19(0.37) | 0.51 | 0.609 |  |
| AgeC | -0.30(0.02) | -12.45 | **<0.001** |  |
| AgeC^2^ | -0.01(0.00) | -3.63 | **<0.001** |  |

Note: AgeC, mean-centred chronological age; AgeC^2^, mean-centred chronological age squared; HC, healthy controls; CI, confidence intervals; SAD, social anxiety disorder; SE, standard error. 𝛃 is measured in years.

|  | N | N AG | N PD | N GAD | N SPH | N Other Anx | % Female | Mean (SD) age |
| --- | --- | --- | --- | --- | --- | --- | --- | --- |
| BCM/MIND-MB | 19 | 1 current | 1 current | 17 current | 1 current | 1 current | 47.40 | 28.11 (6.99) |
| Chiba | 4 | 3 current | 0 | 3 current | 1 current | 0 | 25.00 | 31.75 (9.39) |
| Columbia MRT | 1 | 0 | 0 | 1 current | 0 | 0 | 100.00 | 26.00 (0.00) |
| Columbia SAD | 3 | NA | 0 | 3 current | 1 current | 0 | 33.30 | 33.33 (15.50) |
| Columbia SPP | 5 | NA | 0 | 2 lifetime | 4 lifetime | 0 | 100.00 | 37.40 (11.59) |
| DCCN | 3 | 1 lifetime; 1 current | 1 current | 1 current | NA | 0 | 100.00 | 27.33 (5.13) |
| Dresden | 4 | 1 current | 2 current | 0 | 1 lifetime; 2 current | 0 | 100.00 | 24.75 (4.86) |
| FOR2107 MR | 11 | 1 lifetime; 5 current | 1 lifetime; 4 current | 3 current | 1 current | 0 | 81.80 | 30.82 (6.78) |
| FOR2107 MS | 10 | 1 lifetime; 2 current | 2 lifetime; 1 current | 3 current | 2 lifetime; 1 current | 0 | 50.00 | 40.00 (14.34) |
| Houston | 5 | 2 current | 4 current | 0 | 2 current | 0 | 40.00 | 36.80 (6.91) |
| Istanbul | 2 | 0 | 0 | 0 | 2 current | 0 | 100.00 | 36.50 (10.61) |
| LFLSAD | 4 | 1 current | 1 lifetime; 2 current | 1 current | 1 current | 0 | 75.00 | 44.00 (6.63) |
| MSAD | 5 | 3 current | 3 current | 1 current | 1 current | 0 | 60.00 | 38.00 (13.78) |
| NESDA Amsterdam | 25 | 15 current | 13 current | 2 lifetime; 12 current | NA | NA | 56.00 | 38.60 (10.15) |
| NESDA Leiden | 26 | 4 lifetime; 12 current | 3 lifetime; 13 current | 3 lifetime; 10 current | NA | NA | 65.40 | 41.15 (8.42) |
| NESDA Groningen | 28 | 1 lifetime; 17 current | 2 lifetime; 18 current | 4 lifetime; 11 current | NA | NA | 75.00 | 38.32 (9.35) |
| SP Muenster | 9 | 0 | 2 lifetime; 3 current | 0 | 3 lifetime; 1 current | 0 | 88.90 | 40.33 (9.19) |
| Umea Vox | 7 | 3 lifetime | 2 lifetime; 3 current | 2 current | NA | NA | 71.40 | 30.86 (7.80) |
| Seoul SAD AC | 6 | 1 lifetime; 3 current |  | 0 | 2 current | 0 | 50.00 | 26.17 (3.43) |
| YSAD | 7 | 0 | 1 current | 0 | 0 | 0 | 57.10 | 22.86 (0.90) |
| Total across samples | 184 | 12 lifetime; 66 current | 13 lifetime; 69 current | 11 lifetime; 68 current | 10 lifetime; 16 current | 1 current | 65.20% | 35.44 (10.31) |

Table S10. Sociodemographic information for SAD participants included in SAD (ANX comorbidity) sub-analysis.

Note. AG, agoraphobia; Anx, anxiety; GAD, generalised anxiety disorder; N, number; NA, not applicable; PD, panic disorder; SAD, social anxiety disorder; SD, standard deviation; SPH, specific phobia

Table S11. Sociodemographic information for SAD participants included in SAD (no ANX comorbidity) sub-analysis.

|  | N | % Female | Mean (SD) age |
| --- | --- | --- | --- |
| BCM | 23 | 30.40 | 36.90 (12.80) |
| Chiba | 11 | 54.60 | 32.40 (9.36) |
| Columbia MRT | 25 | 44.00 | 28.00 (6.09) |
| Columbia SAD | 12 | 66.70 | 29.60 (7.18) |
| Columbia SPP | 11 | 72.70 | 33.30 (6.54) |
| DCCN | 12 | 25.00 | 35.80 (9.83) |
| DelMar | 24 | 83.30 | 26.30 (7.18) |
| Dresden | 8 | 37.50 | 29.00 (5.50) |
| FOR2107 MR | 13 | 69.20 | 33.90 (10.20) |
| FOR2107 MS | 13 | 69.20 | 43.60 (9.07) |
| Houston | 15 | 46.70 | 40.30 (12.00) |
| Istanbul | 22 | 27.30 | 33.20 (6.69) |
| LFLSAD | 6 | 66.70 | 46.70 (2.73) |
| UC Louvain | 16 | 100.00 | 23.90 (2.67) |
| LUMC | 15 | 33.30 | 31.50 (7.15) |
| MSAD | 12 | 33.30 | 38.00 (15.00) |
| NESDA Amsterdam | 9 | 44.40 | 37.70 (8.40) |
| NESDA Leiden | 5 | 80.00 | 27.20 (3.03) |
| NESDA Groningen | 5 | 60.00 | 37.80 (9.04) |
| SP Muenster | 50 | 78.00 | 32.90 (9.96) |
| TIP | 9 | 55.60 | 26.20 (3.35) |
| UCSD Sapient Insula | 16 | 43.80 | 29.90 (8.83) |
| Umea Sofie | 21 | 90.50 | 33.90 (8.90) |
| Umea Vox | 12 | 66.70 | 29.80 (6.22) |
| Seoul | 22 | 45.40 | 25.70 (2.36) |
| YSAD | 5 | 40.00 | 22.80 (0.84) |
| Total across samples | 392 | 57.90 | 32.20 (9.68) |

Note. N, number; NA, SAD, social anxiety disorder; SD, standard deviation.

**Relationship Between Clinical Variables in SAD Subgroups**

Table S12. Sample overlap between SAD participant subgroups based on comorbidity and medication use.

| Subgroup | N | % of sample | % Med use | % MDD comorbidity | Mean (SD) LSAS | Mean (SD) STAI-T |
| --- | --- | --- | --- | --- | --- | --- |
| SAD (medication use) | 163 | 28.30 | - | 79.14 | 76.42(23.77) | 60.27(8.73) |
| SAD (no medication use) | 399 | 69.27 | - | 36.09 | 73.53(21.47) | 50.02(10.91) |
| SAD (ANX comorbidity) | 184 | 31.94 | 44.02 | 76.63 | 75.50(22.39) | 56.00(11.46) |
| SAD (no ANX comorbidity) | 392 | 68.06 | 20.92 | 35.46 | 73.57(21.57) | 51.63(11.05) |
| SAD (MDD comorbidity) | 280 | 48.61 | 46.07 | - | 72.09(22.96) | 57.63(10.09) |
| SAD (no MDD comorbidity) | 296 | 51.39 | 11.49 | - | 74.50(21.23) | 47.95(10.38) |

Note: ANX, anxiety; LSAS, Liebowitz Social Anxiety Scale; Med, medication; MDD, major depressive disorder; n, number; SAD, social anxiety disorder; SE, standard error; STAI-T, State Trait Anxiety Inventory – Trait. Data not available for all SAD participants within each group for LSAS and STAI-T. % medication use and MDD comorbidity is calculated within each subgroup.

**Sensitivity Analysis**

**Primary Linear Mixed-Effects Model Results, Removal of n=3 NESDA Sites**

Table S13. Between-group differences in brain-PAD, SAD with ANX comorbidity (n=105) vs HCs (n=1298), exclusion of NESDA sites.

|  | 𝛃 (SE) | t-value | *p* |
| --- | --- | --- | --- |
| Intercept (site) | 0.96(0.76) | 1.27 | 0.206 |
| Diagnosis | 1.92(0.70) | 2.74 | **0.006** |
| Sex | 0.04(0.37) | 0.10 | 0.923 |
| AgeC | -0.32(0.02) | -14.16 | **<0.001** |
| AgeC^2^ | -4.32 x 10^-3^ (1.67 x 10^-3^) | -2.58 | **0.010** |

Note: AgeC, mean-centred chronological age; AgeC^2^, mean-centred chronological age squared; HC, healthy controls; CI, confidence intervals; SAD, social anxiety disorder; SE, standard error. 𝛃 is measured in years.

**Post-Hoc Sensitivity Analysis**

**Primary Linear Mixed-Effects Model Results in SAD with Anxiety Comorbidities and HCs, Removal of Comorbidities One at a Time**

Table S14. Between group differences in brain-PAD: SAD with ANX comorbidities, removal of specific diagnoses.

|  | SAD | HC | Dx | | |  |
| --- | --- | --- | --- | --- | --- | --- |
|  | **n** | **n** | 𝛃 **(SE)** | **t-value** | ***p*** | **Cohen’s *d* (95% CI)** |
| SAD with ANX comorbidity (no GAD) vs HCs | 98 | 1355 | 2.88(0.76) | 3.80 | **<0.001** | 0.33(0.229-0.436) |
| SAD with ANX comorbidity (no PD) vs HCs | 102 | 1355 | 2.65(0.73) | 3.65 | **<0.001** | 0.32(0.21-0.0.42) |
| SAD with ANX comorbidity (no AG) vs HCs | 106 | 1355 | 1.72(0.72) | 2.40 | **0.017** | 0.20(0.10-0.31) |
| SAD with ANX comorbidity (no SPH) vs HCs | 158 | 1355 | 1.72(0.66) | 2.62 | **0.009** | 0.18(0.08-0.28) |

Note: ANX, anxiety; CI, confidence intervals; FDR, false discovery rate; HC, healthy controls; n, number; SAD, social anxiety disorder; SE, standard error. 𝛃 is measured in years
